# ImputeBench: Benchmarking Single Imputation Methods

**DOI:** 10.1101/2025.02.02.25321536

**Authors:** Robin Richter, Juliana F. Tavares, Anne Miloschewski, Monique M. B. Breteler, Sach Mukherjee

## Abstract

Biomedical data often contain missing values and in many applications missing value imputation (MVI) is an important part of the data analysis work-flow. However, the performance of MVI methods depends on details of the joint distribution of data and missingness patterns that are typically unknown in practice, making an *a priori* choice of MVI method challenging. Furthermore, technical assumptions underlying MVI methods can be hard to directly verify in practice. Motivated by these issues, in this paper, we propose an approach for the context-specific selection of MVI methods. Due to the fact that different methods may work well in different cases we argue for a move away from a “one size fits all” view and put forward in this paper a standardized, empirical approach in which MVI methods are benchmarked in the specific context of a problem of interest. We connect our work to the large body of MVI research, along the way refining definitions of missing at random and missing not at random and providing a detailed review of existing work on benchmarking. Our approach can be tailored to reflect specific assumptions on missingness patterns, allowing for application in diverse applied problems. Furthermore, in addition to using real data, we study benchmarking via data simulation spanning a broad range of properties, such as latent factors, non-linearity and multi-modality, with interpretable simulation parameters that are amenable to user specification. The approaches we propose can be used to (i) select an MVI method for a given data set or (ii) benchmark a novel MVI method across a range of regimes. Alongside the general protocol, we provide a specific, reproducible implementation (in the R-package `ImputeBench`, available under github.com/richterrob/ImputeBench) that gives users a ready-to-use tool for MVI selection and assessment. We illustrate the use of `ImputeBench` to study the behaviour of a range of existing imputation methods (k-nn, soft impute, missForest, MICE) in the context of real data from an ongoing large-scale population-level study.

## 1 Introduction

Missing values are a common issue in a broad range of biomedical data analyses [see, among many others, 65, 45, 58]. A simple approach to deal with missing values involves considering only samples with no missing entries – known as *complete case analysis* (CCA, also known as *list-wise deletion*) – however this not only loses information but may introduce bias, for details see the discussions in [69, 74, 23, 43]. Instead, various missing value imputation (MVI) methods are commonly used, that take in incomplete datasets as input and produce a completed version by filling in “plausible” values for the missing entries. For discussion of the appropriateness of CCA or MVI under different assumptions consider for example [13, 5, 30].

The diversity of MVI methods in the literature is large, stemming not only from the fact that the problem is prevalent in many applied fields, but also from the fact that under different data-missingness regimes (i.e. different joint distributions over the data and missingness patterns) different methods may be appropriate. For a recent, broad review of MVI methods we refer the reader to [50] and to Appendix A of the Supplementary Materials, meanwhile, benchmarking papers comparing MVI methods include [9, 97, 15, 93, 45, 94]. There are numerous benchmarking papers in the literature for MVI methods, due to time- and space-constraints however, they are often limited in scope, featuring a limited set of MVI methods and/or a limited range of data-missingness distributions. Furthermore, in practice details of the underlying data-missingness distribution that would be needed for an entirely *a priori* choice of method based on theory is usually unavailable. As a result of these factors, for the practitioner choosing an MVI method appropriate for the data at hand is often a non-trivial problem.

Motivated by the foregoing, in this paper we argue in favour of benchmarking MVI methods on a case-by-case basis. To this end, we propose a standardized benchmarking protocol called *ImputeBench*, and, a specific, reproducible implementation of this protocol as the R-package `ImputeBench`, which entails proposed data and missingness pattern simulation protocols. To the best of the authors’ knowledge such a protocol has not been proposed before. We aim to provide a inclusive view of the topic, targeting a broad audience, from developers of MVI methods to practitioners applying MVI methods. To this end, the paper covers several aspects of the questions of interest, organized into sections, not all of which are aimed at all readers to the same extent. In Section 2 we give a detailed overview of the contributions of this paper and a guide on how to read the paper depending on background and needs. The specific contributions of this paper are:

- Section 3: Refined definitions of *missing at random* (MAR) and *missing not at random* (MNAR) – called *column MAR* and *column MNAR*, respectively – as needed to specify a case-by-case benchmarking protocol.
- Section 4: A review of benchmarking papers on MVI methods.
- Section 5: The benchmarking protocol ImputeBench. This is a conceptual approach that could be implemented/realized in various ways.
- Section 6: A specific implementation of ImputeBench in the R-package `ImputeBench`, including:
  – Section 6.1: A novel data simulation protocol.
  – Section 6.2: A novel missingness pattern simulation protocol.
- Section 7.2: A work-flow detailing how to conduct MVI benchmarking via `ImputeBench`.

A key novelty of the ImputeBench protocol is that it standardizes the bench-marking of MVI methods. This allows both for selection of an MVI method for a given data set or for the study of a novel MVI method on simulated data. Furthermore, we instantiate the general idea by proposing a specific protocol (for simulating test data and a tailored simulation protocol for simulating missing patterns using either simulated or real data) implemented in the R-package `ImputeBench`. This provides a ready-to-use *tool* for user-friendly, reproducible benchmarking of MVI methods. The attempt at standardization of MVI bench-marking provided in this paper is in a similar spirit as the R-package `MixSim` of [57], which proposes a data simulation protocol to benchmark clustering algorithms. The distinction between the benchmarking protocol ImputeBench and the package `ImputeBench` is necessary as the former is a general scheme (that could be implemented in various ways) while the latter is a very specific implementation which, by its nature, makes a number of particular design choices. The guiding idea for the data and missingness pattern protocols of `ImputeBench` is balancing maximal flexibility in the generated data and missingness patterns with a minimal number of (interpretable) parameters.

The primary use-case of `ImputeBench` is in the analysis work-flow for datasets with missing values, in the spirit of the guidelines proposed in [15, 48, 11, 63]. The motivating concern is that due to the subtle way in which MVI performance depends on underlying factors (we show many examples below) even a seemingly sensible choice of MVI method may in fact be highly suboptimal. Hence, before an MVI method is employed, we suggest to use `ImputeBench` to benchmark a selection of MVI methods using the actual data at hand. The idea is to support an informed choice by transparently showing how various approaches perform under circumstances that are plausible for the data at hand. As an aid to practical use, we include also a detailed work-flow on how to benchmark MVI methods with `ImputeBench`, in particular setting out how to specify appropriate missingness scenarios.

A second use-case for `ImputeBench` is for developers of methodology, specifically to test a proposed novel MVI method on simulated data with a wide range of possible underlying distributions. This provides researchers a standardized way to understand – under practical, finite sample conditions – how a proposed new method may add to the existing MVI tool kit under different data-missingness distributions. We note that MVI benchmarking on simulated data is important in its own right, since comparing performance only on real, complete cases might lead to biased results due to bias in the observed data. Compare for example [33] in which phylogenetic data is simulated in order to benchmark MVI methods.

`ImputeBench` is designed to evaluate *single* MVI methods opposed to *multiple* MVI methods, which have been an important strand in the literature. While single MVI methods return from given observed data **X**_obs_ a *single* imputed complete data set 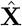, multiple MVI methods yield a set 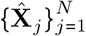 of *N* different complete data sets, imputing for a missing value a vector or a distribution of “plausible” values. Multiple MVI methods are designed to incorporate uncertainty and improve subsequent estimation, testing and regression performance, see [70, 71] and an introduction to multiple MVI methods by [89]. Multiple MVI methods can be used as single imputation methods by choosing an appropriate summary statistic on the vector or distribution of imputed values. As evaluation of multiple MVI methods requires studying performance on subsequent downstream tasks (such as estimation and prediction), at this point it would exceed the scope of this paper, hence we restrict attention to single imputation methods (we discuss extension to the multiple MVI case in Discussion below). The paper is structured as follows. Section 2 contains short summaries of the contributions of this paper together with a guide on where to find relevant information for the respective target audiences. Section 3 gives an overview of common missingness mechanisms including an extension by the introduction of *column MAR* and *column MNAR*. Section 4 provides a detailed review of MVI benchmarking, underlining the need for a *benchmarking tool*. The ImputeBench benchmarking protocol is proposed in Section 5. Section 6 is a detailed account of the proposed data simulation protocol, the missingness pattern simulation protocol and the remaining choices made in the specific implementation of ImputeBench as the benchmarking R-package `ImputeBench`. In Section 7.1 we illustrate the benchmarking of several existing MVI methods – k-nearest neighbour imputation (k-nn), multiple imputation by chained equations (MICE), missForest and soft impute – using `ImputeBench` on simulated data. Section 7.2 sets out the `ImputeBench` work-flow for MVI benchmarking and illustrates the workflow with a real data example involving nutritional data from an ongoing population-level phenotyping study. We conclude with a summary and possible extensions of this work in Section 8.

## 2 Contributions and Guide for the Reader

This section provides a summary of contributions and a guide for readers aimed at efficiently finding relevant Sections of the paper to address specific questions/needs. Depending on interests and needs not all Sections will be of equal interest, hence the suggestions below.

### Guide for the Reader

- *Given a data set with missing entries, how to conduct MVI benchmarking with* `ImputeBench`*?* Primary: Section 7.2; Secondary: Sections 5, 6.2 and 6.3 and Appendix A of the Supplementary Materials.
- *Given a data set with missing entries, how does* `ImputeBench` *benchmark MVI methods?* Primary: Sections 5, 6.2 and 6.3; Secondary: Sections 7.2 and 3.
- *Given a new MVI method, how does* `ImputeBench` *benchmark MVI methods on simulated data?* Primary: Sections 5, 6.1, 6.2, 6.3 and 7.1; Secondary: Appendices A and B of the Supplementary Materials.
- *How does* `ImputeBench` *define MCAR, MAR and MNAR?* Primary: Section 3; Secondary: Section 6.2.
- *How do MVI methods compare in previous benchmarking studies?* Primary: Section 4; Secondary: Section 7.1 and Appendix A of the Supplementary Materials.

### The Definition of Column MAR and Column MNAR (Section 3)

Missingness patterns are commonly categorized by their underlying *mechanism*, i.e. in what distributional relation they are to the values of the complete data matrix. The common distinction of *missing completely at random* (MCAR), *missing at random* (MAR) and *missing not at random* (MNAR) can be defined as a property of either the underlying distribution or of a specific draw [see e.g. 77]. We argue that for a most useful distinction in practice MCAR, MAR and MNAR should be properties of the variables, i.e. columns of the data matrix. To this end, we introduce *column MAR* and *column MNAR* as properties of the marginal distributions.

### A Review of Benchmarking of MVI Methods (Section 4)

In Tables 1 and 2 we compare 34 benchmarking papers on MVI with regards to the methods they compare, which method was shown to perform best in at least one category, what kind of data they used, what kind of missingness mechanisms they imposed, if they varied any parameters and how they measured performance. We observe a lack of standardization, and, often inconclusive results, which we argue is due to the dependence of relative performance on details of the data-missingness distributions, motivating a need to benchmark MVI methods on a case-by-case, rather than “one-size-fits-all” basis.

**Table 1:**
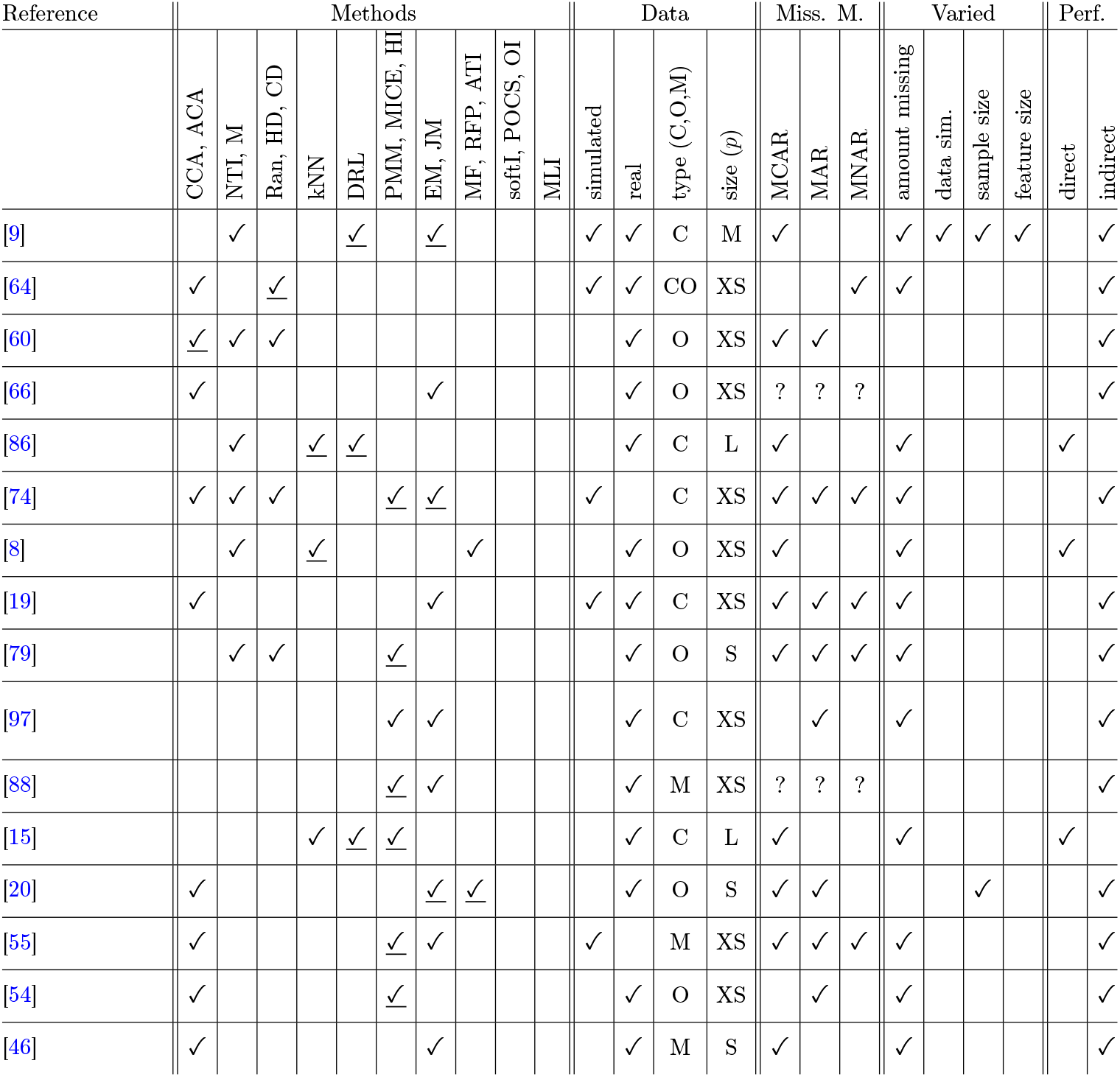
Overview of benchmarking papers of MVI methods (Full details of abbreviations for the MVI methods appears in Appendix A of the Supplementary Materials; Miss. M. = missingness mechanism; Perf. = performance measure; the data types are denoted by C = continuous, O = ordinal/categorical and M = mixed; the data sizes are denoted by XS = *p* ≤ 15, S = *p* ∼ 15 − 80, M = *p* ∼ 80 − 900, L = *p* ∼ 900 − 9000; underlined <u>✓</u> indicates that the paper suggest one of these methods to be superior at least in one tested scenario; three ?’s in the missingness mechanism columns signify that real missing entries were used with unknown underlying mechanism; performance measures are divided into *direct* (e.g. RMSE on imputed data set against a ground truth) and *indirect* (e.g. estimation performance after imputation)).

**Table 2:**
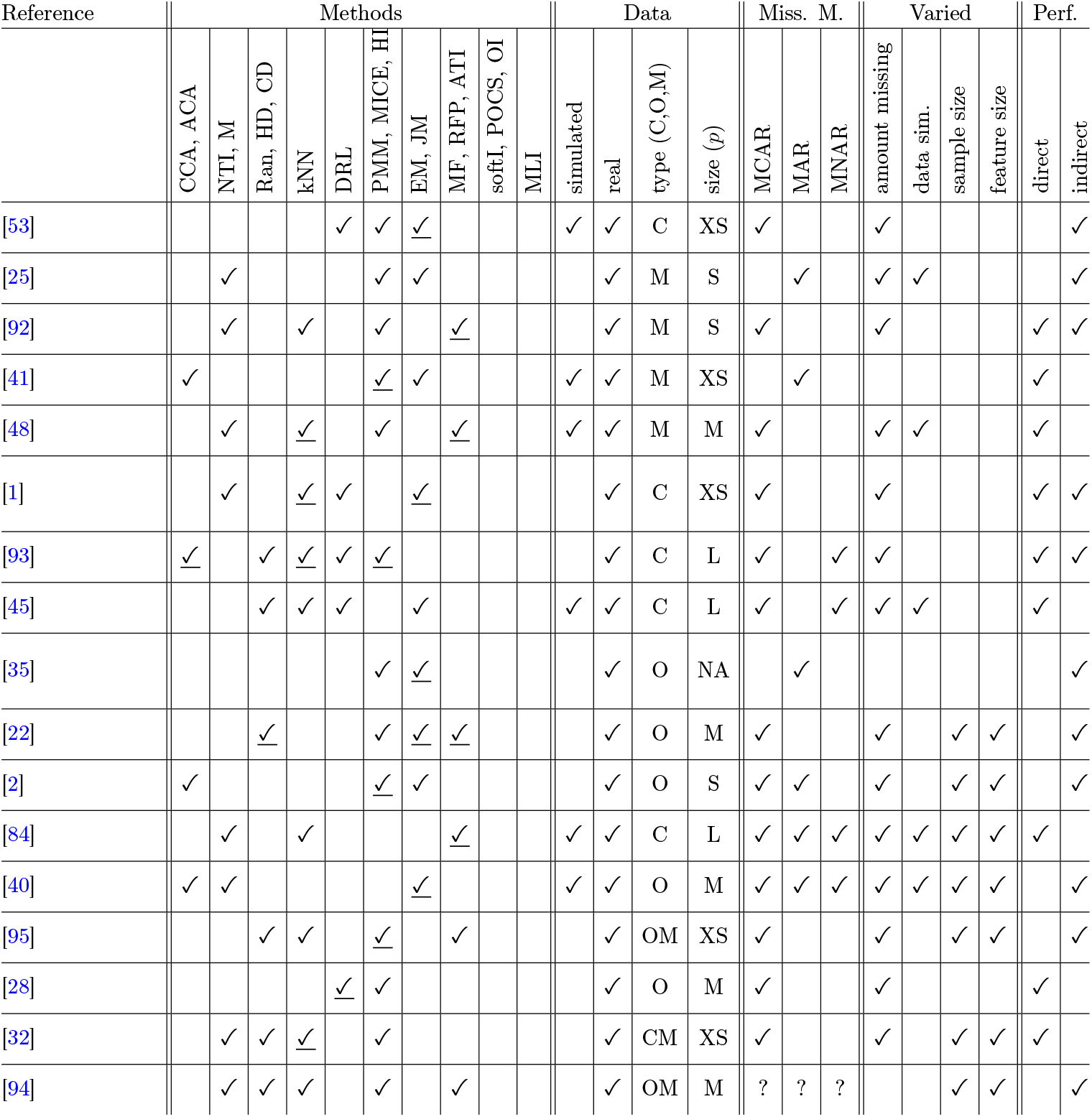
Continuation of Table 1.

### A General Protocol to Benchmark MVI Methods (Section 5)

To perform benchmarking of MVI methods on a case-by-case basis we propose a general benchmarking scheme for real and simulated data, cf. Figure 1. The main idea is to draw for a fixed number of repetitions additional missingness patterns for data that is either given or drawn itself, creating in this way a new observed data matrix in turn imputed by the MVI methods compared and evaluated via an appropriate loss function. For interpretable results we furthermore propose to choose one MVI method to be the “baseline” and the output of the comparison to be the empirical distribution of the relative performances of the MVI methods with respect to the baseline MVI method.

**Figure 1:**
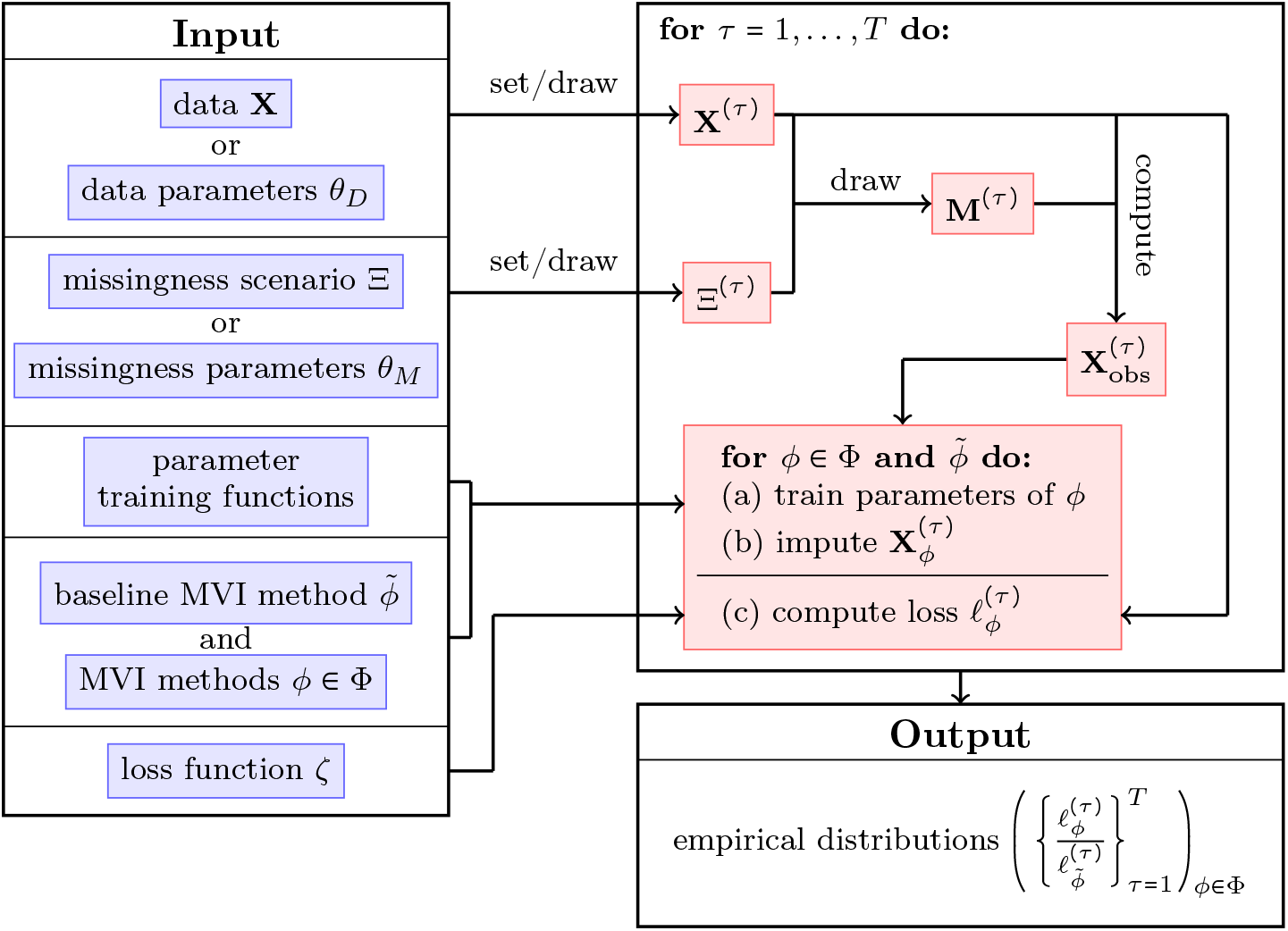
Schematic overview of the ImputeBench protocol.

### A Specific Data and Missingness Pattern Simulation Protocol for the ImputeBench R-package (Section 6)

First, we propose a data simulation protocol guided by balancing the flexibility of the drawn data with respect to the inclusion of data properties observed in real data, and, the interpretability and amount of tuning parameters. The proposed data simulation protocol, cf. Figure 3, allows among others for mixed data, latent low-dimensionality, non-linearity, heavy-tailedness and multi-modality in the marginal distributions. To the end of providing a way to steer a missingness pattern simulation, we define a *missingness scenario* that allows the user to employ tailored assumptions on the missingness pattern for the MVI benchmarking protocol. Last, we provide more details on the design choices of `ImputeBench`.

### A Work-flow on How to Use ImputeBench on Real Data (Section 7.2)

A suggested work-flow for MVI benchmarking on real data is presented, detailing how missingness scenarios might be chosen, what other choices in `ImputeBench` should be considered and how the imputed data might be visually checked. This is illustrated in the context of nutritional data from an ongoing population-level phenotyping study.

## 3 Missingness Mechanisms

Three types of missingness mechanisms are commonly distinguished by independence relations of the joint distribution of the data and the missingness pattern: MCAR (missing completely at random), MAR (missing at random) and MNAR (missing not at random; sometimes known as NMAR). This section covers their respective definitions in the literature and offers a refinement for the MAR and MNAR mechanism called *column MAR* and *column MNAR*, respectively, that will be useful in defining simulation protocols.

Considered in the following are a *latent data matrix* **X**_lat_ ∈ ℝ^*n*×*p*^, a binary *missingness mask* **M** ∈ {0, 1} ^*n*× *p*^ and an *observed data matrix* **X**_obs_ ∈ (ℝ ∪ {NA})^*n*×*p*^, where NA denotes a missing entry, with

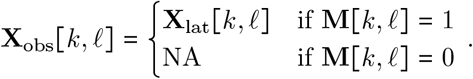

Note that for the sake of simplicity of exposition we define here an entry **X**_lat_ [*k, ℓ*] to be real, however, there are many instances of missing values in categorical/ordinal or binary data.

Which entries of **X**_obs_ to impute and if so which imputation method to use on which entries depends on the joint distribution of (**M, X**_lat_), as it determines the distribution of **X**_obs_ and in turn any discrepancy in analysis results using an imputed or CCA-reduced matrix version of **X**_obs_ in comparison with using the latent **X**_lat_. As is common, we assume in the following that the rows/samples of **X**_lat_ and **M** are given as *iid* draws

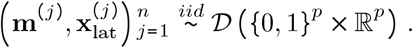

The most straightforward missingness mechanism is *MCAR* as it assumes the distribution of values in **M** to be independent of **X**_lat_.

### Definition 3.1

(e.g. [52]). Let **m**, (**x**_lat_) ∼ 𝒟 and ℙ [**m [***k*] = 0] ≠ 0 for some 1 ≤ *k* ≤ *p*, we call the *k*-th entry in **x**_obs_ *missing completely at random* (MCAR) if for any distinct vectors **x**_0_, **x**_1_ ∈ ℝ^*p*^ we have

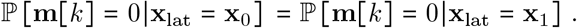

Meanwhile, the missingness mechanism *MAR* is defined as the case that **M** only depends on *observed* values of **X**_lat_, i.e. values present in **X**_obs_, the idea being that the information as to why a value is missing is present in **X**_obs_ itself. Defining this as a property of 𝒟 is however not straight-forward and definitions in the literature have not been consistent, see for a discussion on this issue [77], whose distinction in *realized* and *everywhere* MAR we pick up in the following.

### Definition 3.2

([77]). Let 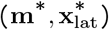 be a realization of (**m, x**_lat_) ∼ 𝒟 and let **m**^*^ [*k*] = 0 for some 1 ≤ *k* ≤ *p*. Given a vector **y** ∈ ℝ, denote in the following by **y**_(−*k*)_ ∈ ℝ^*p*−1^ the same vector without the *k*-th entry. We say the entry 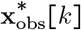 is *realized missing at random* (realized MAR) if for any distinct vectors x_0_ ∈ ℝ^*p*^ with **x**_0_ [*ℓ*] = **x** [*ℓ*]for all [*ℓ*] ∈ {*r* : **m**^*^ [*r*] = 1 and *k* ≠ *r*} we have

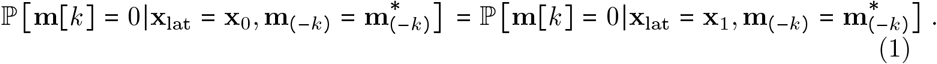

In contrast to realized MAR being defined above as a property of a realization of (**M, X**_lat_) the following definition defines MAR as a property of the underlying distribution *𝒟*.

### Definition 3.3

([77]). Let (**m, x**_lat_) 𝒟∼ and ℙ [**m** [*k*] = 0 ≠ 0 for some 1 ≤ *k* ≤ *p*, we call the *k*-the entry in **x**_obs_ *everywhere missing at random* (everywhere MAR) if *for any* **y** ∈ {0, 1} ^*p*^ s.t. ℙ [**m** = **y**] ≠ 0 and for any distinct vectors **x**_0_, **x**_1_ ∈ ℝ^*p*^ s.t. **x**_0_[*ℓ*] = **x**_1_[*ℓ*] for all *ℓ* ∈ {*r* : **y**[*r*] = 1 and *k* ≠ *r*} we have

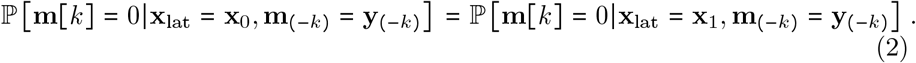

Note that the definitions of MCAR and MAR, as they can be found in [52] or [77] are given commonly as characterizations of the *entire* missingness pattern **M** or its underlying distribution. Meanwhile, above we gave a *column-wise* definition of MCAR and MAR, as, distinction between missingness mechanisms per variable is practically more sensible (to take a practical example: in nutritional data, missingness patterns for the variable “alcohol consumption” may be quite different from “tea consumption”, say). Moreover, in our case the columnwise definition of missingness mechanisms is paramount to define a missingness pattern simulation that can mix different missingness mechanisms as later done in Section 6.2. Furthermore, we argue that the definition of everywhere MAR given in Definition 3.3 seems unnecessarily strict on the distribution 𝒟. Giving an example, consider the case that only the first column in **X**_obs_ features missing entries, whose missingness pattern depends exclusively on values of the second column in **X**_obs_. Under Definition 3.3 this would mean missing values of the first column of **X**_obs_ are missing everywhere MAR. Consider now adding missingness in the second column, given that (2) is only satisfied if the second column is observed, then the missing values of the first column would no longer be everywhere MAR, which seems unsatisfactory since the mechanism by which the missingness pattern of the first column is “drawn” did not change. To the end of allowing for such scenarios to “remain” MAR, we propose here a relaxed version of everywhere MAR, which, rather than demanding MAR missingness (1) for any potential missingness pattern **y**, demands that there *exists* a missingness pattern with non-zero probability such that (1) holds.

### Definition 3.4.

Let **m, x**_lat_ ∼ 𝒟and ℙ [**m** [*k*] = 0] ≠ 0 for some 1 ≤ *k* ≤ *p*, we call the *k*-the entry in **x**_obs_ *column missing at random* (column MAR) if there exists a subset *K* ⊆ {1, …, *k* − 1, *k* + 1, …, *p*} such that for any distinct vectors **x**_0_, **x**_1_ ∈ ℝ^*p*^ agreeing on *K*, i.e. **x**_0_[*t*] = **x**_1_[*t*] for all *t* ∈ *K*, we have

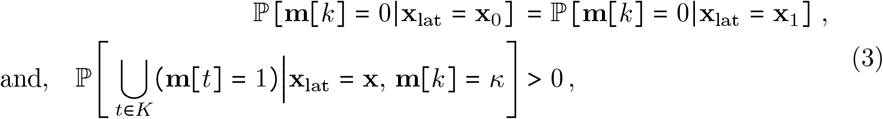

for all *κ* ∈ {0, 1} and **x** ∈ ℝ^*p*^ s.t. ℙ **x**_lat_ = **x** > 0.

Note, that both Equations (2) and (3) are <u>not</u> satisfied as soon as the probability of being missing depends on the (latent) value itself, which is conceptually a necessary requirement for MAR. Moreover, any MCAR value is also missing realized, everywhere and column MAR by construction.

The third missingness mechanism MNAR is commonly defined as the complement of MAR missingness. Being the opposite of MAR, the definition of MNAR values aims at describing the case that **M** depends on values of **X**_lat_ not observed in **X**_obs_. In this paper we work only with column MAR and its counterpart column MNAR, and, for simplicity we omit the term “column” in the following, consequently we will only give the definition for column MNAR and omit realized and everywhere MNAR.

### Definition 3.5.

Let (**m, x**_lat_) ∼ 𝒟 and ℙ **m** *k* = 0 ≠ 0 for some 1 ≤ *k* ≤ *p*, we call the *k*-th entry in **x**_obs_ *column missing not at random* (column MNAR) if there exists no subset *K* ⊂ {1, …, *k* − 1, *k* + 1, *p*} such that for any **x**_0_, **x**_1_ ∈ ℝ^*p*^ agreeing on *K* we have that (2) holds.

While realistic examples of truly MAR missingness may be hard to construct, a prominent case of MNAR missing values is given by missing values in experimental measurements that suffer from a limit of detection.

Note that, distinguishing MCAR, MAR and MNAR missingness is of particular use in understanding the limitations of complete case analysis. In the case of MCAR missingness in a given column the observed entries are still an *iid* sample of its respective latent marginal distribution, meanwhile in the classic case of MNAR missingness – the distribution of missing entries being dependent on the values of its column – the observed entries constitute a *biased* sample of the latent marginal distribution. Meanwhile in the case of MAR missingness it depends on the dependency structure of **X**_lat_ whether or not the observed sample is biased. Note however, while the distinction of MCAR, MAR and MNAR might be a good primer when it comes to complete case analysis vs. MVI methods choosing the “right” imputation methods depends also strongly on the correct modelling assumptions on the dependency structure of **X**_lat_, which is not captured in the definitions of MCAR/MAR/MNAR.

## 4 Benchmarking Papers on MVI

Benchmarking papers provide important guidance for practitioners on which method to employ for the respective data at hand. In MVI benchmarking the large number of methods and even larger number of relevant data-missingness distributions to test on make it necessary to target often very specific settings. An overview of 34 MVI benchmarking papers is given in Tables 1 and 2, the papers are compared by the kind of MVI methods included, the types of test data set(s) considered and its/their size, whether the test data was real or simulated, which missingness mechanisms were considered, if parameters were varied and whether the evaluation was given via a direct imputation error (e.g. RMSE) or an indirect error (e.g. via subsequent estimation of regression coefficients). For an overview of MVI methods and their here used shorthand we refer the reader to Appendix A of the Supplementary Materials. We summarize our finding and give more detail in the remainder of the section.

### MVI Methods and Data Considered

As seen in Tables 1 and 2 the MVI methods compared vary widely over the benchmarking papers. Furthermore, a number of papers show no clear “winner”, that is different MVI methods are recommended for different data-missingness distributions tested, while some benchmarking papers feature no clear winner at all.

Validation on real data examples are most common in the benchmark literature, as there are only two examples that *only* use simulated data ([74, 55]). Meanwhile, simulated data sets are used in less than half of the benchmarking papers (11 out of 33). In the case of simulated data, most simulation protocols involve drawing from multivariate normal distributions. The parameters of these multivariate distributions are chosen either generically or as in [55] based on real data, e.g. by setting the mean and the covariance according to estimated values. An exception is [48] who uses also Poisson draws and [40] where exponential random graphs are simulated. While [84] also uses a MVN distribution to draw data values they additionally include a coupled simulation for data and missingness pattern.

### Missingness Mechanisms Included

MCAR is the main missingness mechanism considered (13 of 30 times MCAR is the *only* mechanism considered, and, only 6 times MCAR is not considered at all). However, in practice especially MAR and MNAR mechanisms are of interest as they lead to larger absolute errors when using the baselines (complete case analysis, mean/median/mode imputation, random draw). When MAR/MNAR mechanisms are considered often the exact simulation protocol is missing, hindering reproducibility of results. The inclusion of the specific missingness pattern simulation protocol is of particular interest in the MAR/MNAR case, as opposed to MCAR missingness, which is straight-forward to generate, generating MAR/MNAR missingness comes with many design choices on the regression pattern that are often not obvious.

### Performance Measures Included

Tables 1 and 2 distinguishes papers that measure the performance of MVI methods *directly* by comparing the imputed matrix with a ground truth and those that measure the performance *indirectly* by means of comparing the performance of a subsequent task such as estimation, regression or classification. In particular the benchmarking papers considering multiple imputation approaches (JM, MICE, etc.) evaluated preferably via indirect measures of performance, as the advantages of multiple imputation such as computable confidence intervals can only be taken into account when measured indirectly.

## 5 The ImputeBench Protocol

We argue in favour of a framework for MVI benchmarking that

- allows for benchmarking on real and simulated data,
- can be tailored to specific use-cases,
- outputs robust, reproducible performance comparisons, and,
- yields interpretable results.

To this end, we introduce in this section an MVI benchmarking protocol called *ImputeBench*. In the following section we go on to propose furthermore an implementation of the ImputeBench protocol available in the accompanying R package `ImputeBench`. A high level overview of ImputeBench is given in Figure 1.

### Idea and Nomenclature

The ImputeBench protocol tests MVI methods by applying them to a data matrix with artificially missing entries and subsequent comparison with the ground truth. To have an interpretable measure of loss we propose in ImputeBench to compare each imputation loss with the imputation loss of a baseline approach chosen by the user. To this end, ImputeBench must include data simulation and missingness pattern simulation protocols. First, in the case the user does not provide data, data is generated via the data simulation protocol using *data parameters θ*_*D*_ as input, otherwise the user-provided data is used. Second, the missingness pattern simulation protocol governs how the additional missingness pattern is drawn given data and some formalization of the assumed, underlying missingness mechanisms. To this end, we propose to distinguish *missingness scenarios* Ξ and *missingness parameters θ*_*M*_ . While the former should detail column-wise specific missingness mechanisms, the latter should be a more unspecific vector of parameters from which a missingness scenario is drawn. A missingness scenario aims at giving the practitioner the opportunity to specify very tailored missingness pattern when the goal is to test MVI methods on real data with genuinely missing entries, where very specific assumptions on the origin of the missing entries need to be taken into account (for an example see Section 7.2). Definition 6.3 gives an example of a missingness scenario. Meanwhile, specifying a missingness scenario on simulated data is not practical, hence, the need for the additional definition of missingness parameters and a protocol how to draw missingness scenarios from missingness parameters.

#### Note 5.1.

*The data and missingness pattern simulation protocols and the definition of a missingness scenario are, for the sake of simplicity, not considered inputs to the ImputeBench protocol, cf. Figure 1 (even though technically they are inputs). We note that in practical implementation and use of the ImputeBench protocol (see Section 6) the data and missingness pattern simulation protocols and the definition of a missingness scenario are fixed by context and are thus not user-dependent*.

### ImputeBench Pseudo Code

- <u>Input:</u>
  – Data **X** ∈ (ℝ ∪ {NA})^*n*×*p*^, or, *data parameters θ*_*D*_ ∈ Θ_*D*_.
  – A *missingness scenario* Ξ, for an example see Definition 6.3, or, *missingness parameters θ*_*M*_ ∈ Θ_*M*_ .
  – A loss function ζ : (ℝ ∪ {*NA*}) ^*n*×*p*^ × ℝ^*n*×*p*^ → 0, ∞) serving as performance measure.
  – A baseline MVI method 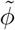.
  – A set of MVI methods Φ.
  – Parameter training functions for the MVI methods (including the baseline method).
  – Number of Repetitions *T* ∈ ℕ.
- <u>Protocol:</u>
  A. **for** *τ* = 1, 2, …, *T* **do:**
    i. **If** data is not provided: Simulate data **X** ^*(τ)*^ ∈ ℝ^*n*×*p*^ according to a data simulation protocol using data parameters *θ*_*D*_, for an example see Section 6.1. **Else:** Set **X** _(_^*τ*)^ = **X**.
    ii. **If** missingness scenario is not provided: Draw missingness scenario Ξ ^*(τ)*^ using missingness parameters *θ*_*M*_, for an example see Section 6.2. **Else:** Set Ξ ^(*τ*)^ = Ξ.
    iii. Simulate missingness pattern **M** ^*(τ*)^ ∈ {0, 1}^*n*×*p*^ according to a missingness pattern simulation protocol using scenario Ξ ^*(τ*)^, for an example see Section 6.2. Denote by **X**_obs_ the data **X**^*(τ*)^ masked by **M**^*(τ*)^.
    iv. **for** each MVI method *ϕ* ∈ Φ and the baseline method 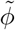 **do:**
      a. Train or set parameters (either only for *τ* = 1 or for each *τ* individually).
      b. Impute 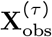 to obtain 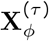
      c. Compute the loss 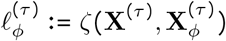.
- <u>Output:</u> The empirical distributions of

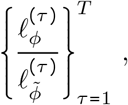

for all MVI methods *ϕ* ∈ Φ.

## 6 The ImputeBench Package

In the following section we propose a data simulation protocol, a notion of *missingness scenario* and a missingness pattern simulation protocol for the ImputeBench benchmarking protocol of Figure 1. The overarching goal of the design choices made for the simulation protocols in this section is balancing the flexibility and broadness of the test data-missingness distributions covered by the resulting R-package `ImputeBench` against the feasibility and interpretability of the user-chosen input parameters *θ*_*D*_ and *θ*_*M*_ . For the sake of completeness we report furthermore the featured default choices of MVI methods, parameter training functions and loss functions of `ImputeBench` in the last subsection of this chapter.

### 6.1 Data Simulation Protocol

The data simulation protocol of `ImputeBench` is designed to include data properties such as mixed dimensionality, latent (low-)dimensionality, (non-)linearity, multi-modality and data type mixture, for examples see Figure 2. We distinguish in the following parameters which control the main properties of the simulated data – called the *primary data parameters* – from *secondary data parameters* that mainly govern the exact shape of the data. For an overview of all parameters see Table 3. We would like to stress at this point, that the goal of this data simulation protocol is not to truly mimic real data – as this may be infeasible in general – but rather to include as many properties found in real data as possible to allow for a wide but still targeted survey of performance characteristics.

**Table 3:**
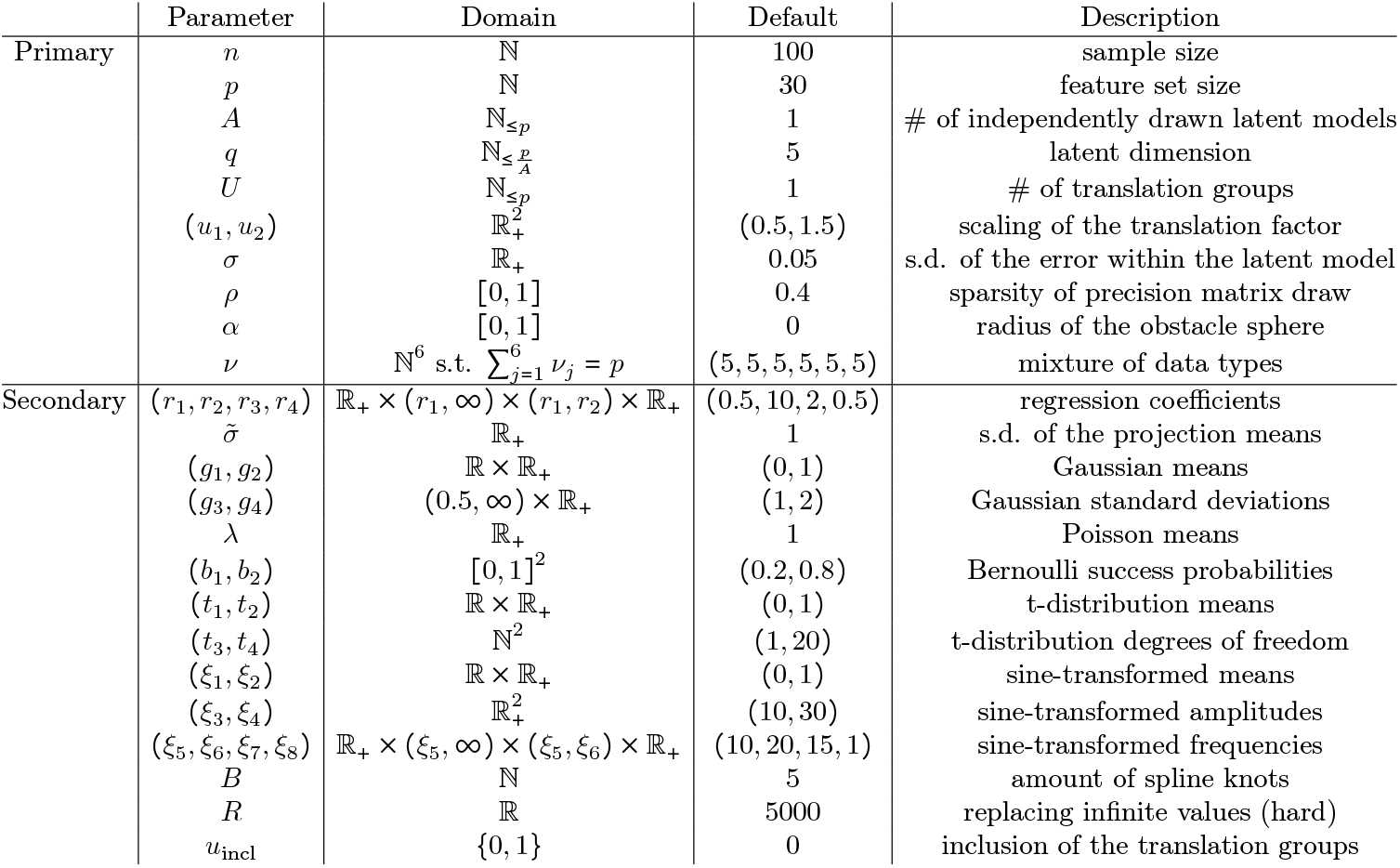
Parameters of the data simulation protocol defining *θ*_*D*_ ∈ Θ_*D*_.

**Figure 2:**
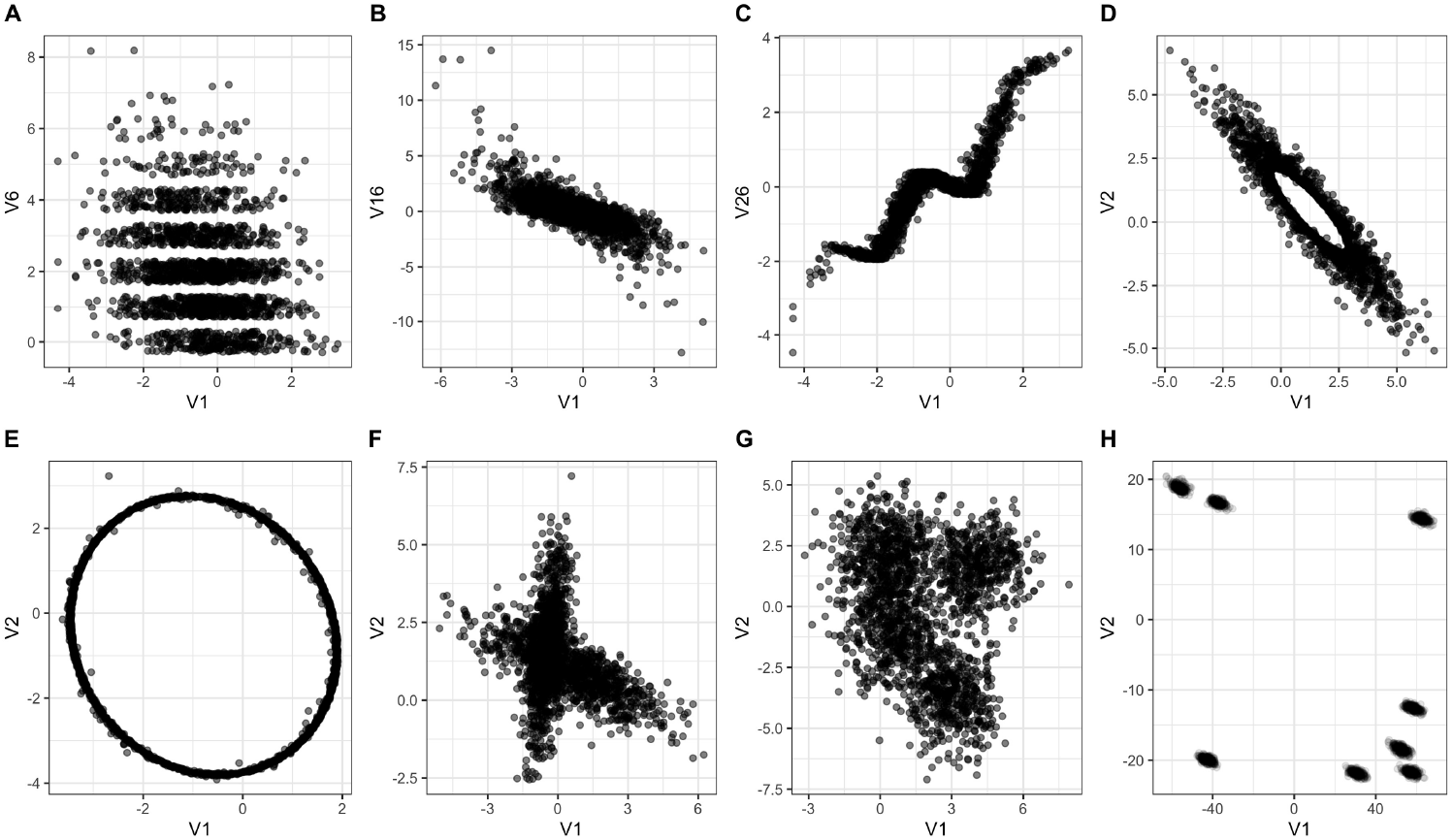
Examples of data simulated with ImputeBench. Exemplified are mixed data (A: normal versus Poisson distributed variable (with added jitter)), heavy-tailedness of marginal distributions (B: normal versus t-distributed variable), non-linearity (C: normal versus spline-transformed variable, D,E: normal variables with “obstacle sphere” (*α* equal to 0.25 and 1, respectively), multiple independently regressed subgroups of samples (F: *A* = 2) and multi-modality in the marginal distributions (G,H: *U* equal to 4 and 8, respectively, with increased scaling of the translation factors in H). Furthermore, all depicted examples follow some kind of latent low-dimensional model.

#### Overview

The data generating protocol is summarized in Figure 3 and consists of two stages. First, in the *MVN-stage* a multivariate normal model is drawn, possibly carrying a latent lower dimensional (non-)linear structure. Second, in the *transformation stage* the outcome of the first stage is (quantile-)transformed to simulate normal, Poisson, Bernoulli and t-distributed variables, as well as, sine and spline-transformed variables, and if chosen, all columns are translated according to a mode assignment. An overview over the data parameter vector *θ*_*D*_ is given in Table 3.

**Figure 3:**
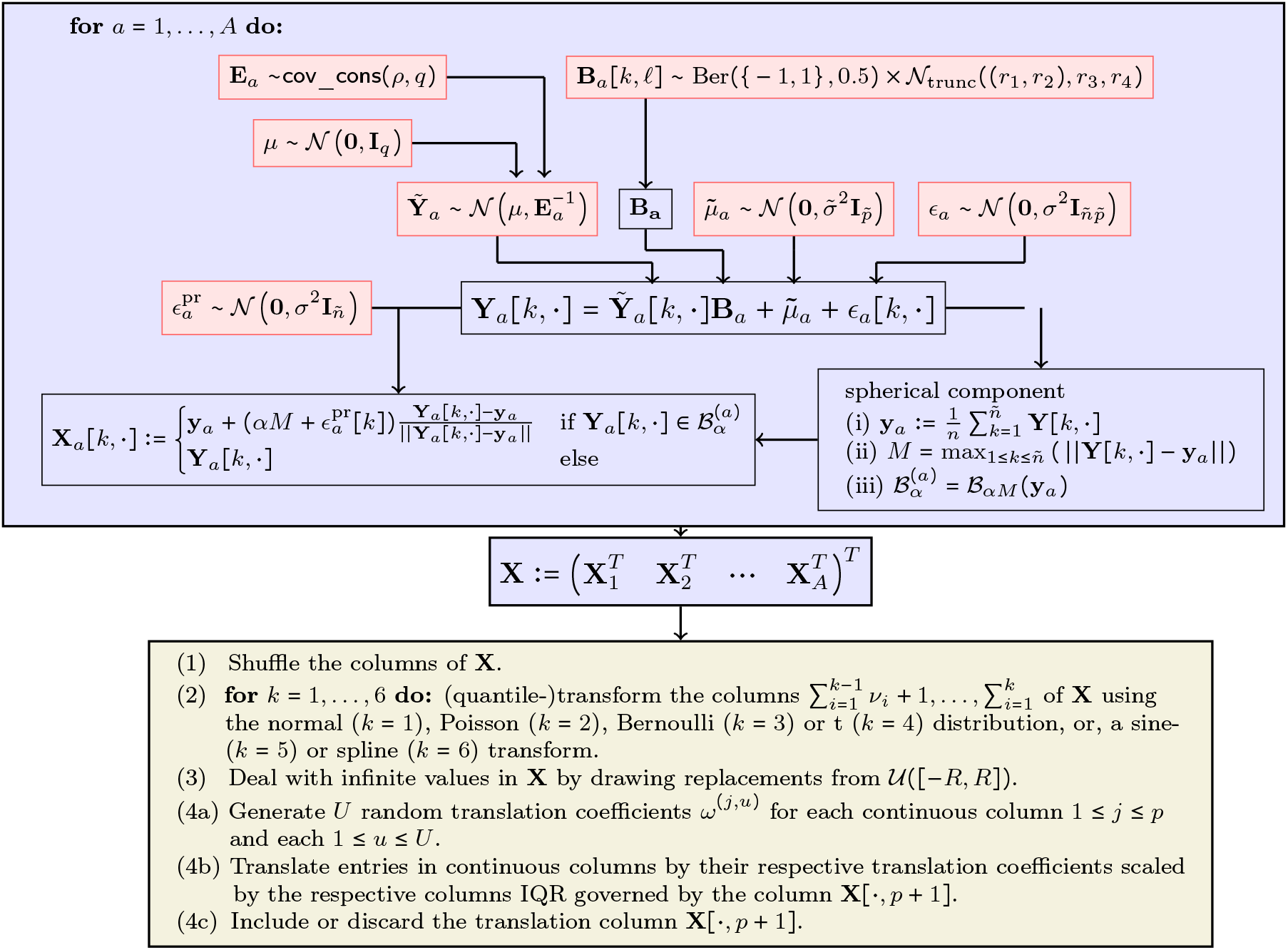
Schematic overview of the data simulation protocol. Depicted in blue are the steps of the *MVN-stage* and in olive the steps of the *transformationstage*.

The overall size of the data is given by *n* ∈ ℕ samples of *p* ∈ ℕ features. Via the six-component vector ν the mixture of data types (Normal/Poisson/Bernoulli/t-distributed/sine-transformed/spline-transformed), enforced in the transformation stage, is defined. Meanwhile, *A* ∈ ℕ is the number of independently drawn latent models in the MVN-stage of size *p* × ^*n*^/_*A*_, each a linear model with latent dimension *q* ≤ *p*, whose latent variables are drawn as a multivariate normal distribution with a precision matrix drawn by the model of the R function cov_cons using Erdös-Renyi games, for details see the R-package genscore ([49, 98]), governed by a sparsity parameter *ρ* ∈ [0, 1] and with additive *iid* normal distributed error terms with standard deviation *σ* ∈ ℝ_+_, cf. the red elements in Figure 3. Hence, *q* governs the latent low-dimensionality of the simulation protocol, and, *A* the option of having overlapping regression models of the same latent dimension, cf. Figure 2 F. Meanwhile, non-linearity is enforced either via the parameter *α* ∈ [0, 1] controlling a spherical structure, cf. Figure 2 D,E, or, in the transformation stage for example by adding spline-transformed columns, cf. Figure 2 C. Last, *U* ≤ *p* is used to control multi-modality in the marginal distributions, determining the number of translation groups which comes into play in the transformation stage, cf. Figure 2 G,H. Last, by the quantile-transformations governed by ν mixed data can be simulated, including features such as ordinal and heavy-tailed variables, cf. Figure 2 A,B.

**MVN-Stage - Details** We set for each group of rows a sample size of

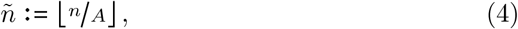

and construct in a first step independent data matrices 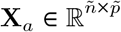, with

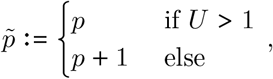

for 1 ≤ *a* ≤ *A*. Recall that previously undefined secondary parameters can be found in Table 3. Note, that for the sake of reducing the number of parameters the data simulation protocol considers only splits into equally sized sample groups via *A*. Find below the pseudo code underlying the draw of **X**_*a*_.

**For** *a* = 1, …, *A* **do:**

1. Draw a precision matrix **E**_*a*_ ∈ ℝ^*q*×*q*^ with set sparsity *ρ* using cov_cons [49, 98].
2. Draw ñsamples from 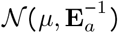 to constitute 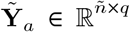, where *µ* ∼𝒩 (0, **I**).
3. Draw a projection matrix 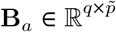 by

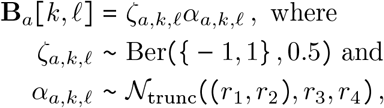

are independently drawn, with Ber denoting the Bernoulli distribution and 𝒩_trunc_ a truncated Gaussian distribution.
4. Draw an error matrix 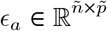 by independently drawing

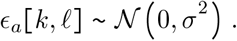
5. Draw 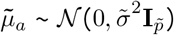 and compute

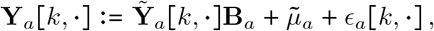

for all 1 ≤ *k* ≤ ñ.
6. Compute the column medians 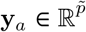 of **Y**_*a*_ and denote by *M* the maximum distance over all rows of **Y**_*a*_ to **y**_*a*_. Then define the obstacle sphere 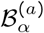 for *α* ∈ [0, 1] as the ball 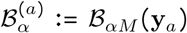 with radius *αM* around **y**_*a*_. Draw

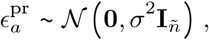

and compute

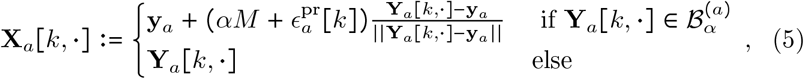

for all rows 1 ≤ *k* ≤ ñ.

##### Remark 6.1.

*To achieve the border case of column-wise independent entries of* **X** *one sets (q, ρ*) = (*p*, 0) *or one lets σ tend to infinity, whilst not adding non-linearity via α* = 0.

Let us elaborate in the following Note on the spherical element introduced in (6.).

##### Note 6.2.

*In the above protocol non-linearity is added via the parameter α* ∈ [0, 1] . *In detail, α is the normalized radius of an* “obstacle sphere”, *used by projecting all points inside the sphere onto its hull, see Figure 2 D*,*E. Hence, via (*5*) the parameter α governs scale-independently and continuous the projection of parts of the linear structure onto a sphere, while keeping the latent dimensionality. In particular, choosing α* = 1 *we obtain a sphere structure of latent dimension q* − 1 *with no linear structure remaining, meanwhile choosing α* = 0 *yields a linear structure of latent dimension q. Note that this “continuous” transition via α from linear to spherical structure is motivated by the generality of the sphere and the absence of additional parameters, rather than actual examples in reality*.

(7.) Combine all independently created data sets to obtain

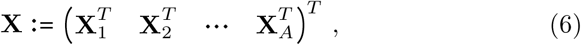

and shuffle the rows. Note that by construction **X** has 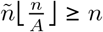 rows.

### Transformation Stage - Details

1. Shuffle and normalize the columns in **X** by subtracting the sample mean and scale by the inverse of the sample standard deviation.
2. For each entry ν_*k*_ consider all columns 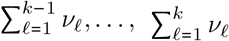 of **X** given they do not constitute the empty set. Then for *k* = 1, 2, 3, 4 quantile-transform the considered columns to Gaussian, Poisson, Bernoulli or t-distributed random variables, respectively, with random parameter choices governed by the secondary parameters in Table 3. The details of these quantile transforms can be found in Appendix B of the Supplementary Materials. For *k* = 5, 6 we use a randomized sine- and spline-transforms, respectively, governed by their respective secondary parameters of Table 3, detailed again in Appendix B of the Supplementary Materials. Last, in case *U* > 1 the column *p* + 1 is quantile-transformed into a discrete uniform distribution on {1, 2, …, *U*}.
3. To ensure robustness all infinite values of **X** are replaced with draws from 𝒰 ([−*R, R*]).
4. **If** *U* > 1 **do:** For each continuous column and each 1 ≤ *u* ≤ *U* a random value scaled by the IQR of the given column is drawn and added to the respective entry of each sample of the respective group.
  a. For each column 1 ≤ *j* ≤ *p* with continuous entries denote by *Z*_*j*_ its respective interquartile distance (IQR). For each group 1 ≤ *u* ≤ *U* draw independently

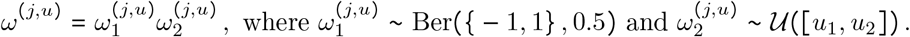
  b. Compute

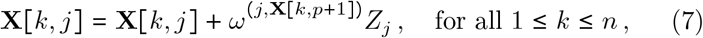

for *j* over all continuous columns. Note, that the primary parameters *u*_1_ and *u*_2_ determine in a continuous fashion how far apart the translation groups are, in particular *u*_1_ = *u*_2_ = 0 would leave the data as is.
  c. **If** *u*_incl_ = 0 **do:** Discard the (*p* + 1) -th column of **X**.

### 6.2 Missingness Pattern Simulation Protocol

As argued in Section 5 in order to implement a missingness pattern simulation in `ImputeBench` we propose a two-fold strategy with the definition of a *missingness scenario* Ξ, giving detailed instruction on the missingness mechanisms of each individual column, and, *missingness parameters* to be used to draw missingness scenario in the case the tailored nature of missingness scenarios are not needed.

In detail, a missingness scenario should

- amend for the possibility of imposing MCAR, MAR and MNAR missingness pattern;
- feature as few parameters as necessary.
- allow for detailed instructions such as “the second column of the missingness pattern should be governed by regression on the fourth column of the data matrix”;

Meanwhile, missingness parameters should be designed with only the first two bullet points in mind.

#### Missingness Scenario

Arguably the most difficult design choice to make when defining a missingness scenario is *how* to impose MAR/MNAR missingness. Hence, before providing a proposed definition of a missingness mechanism, we first spend some time on the choice of regression used in drawing MAR/MNAR missingness. To this end, denote first by **y** ∈ ℝ^*n*^ a score vector that is to be translated into probabilities of being missing. For MAR missingness we compute the score vector **y** as the sum of all standardized chosen regressor columns. The limitation of having all regression coefficients set to 1 is done in order to keep the model simple. Meanwhile, for MNAR missingness **y** is given by the column to be masked itself and no other column is contributing, again this limitation is done for simplicity sake. Note, that by imposing multiple missingness mechanisms on the same column weighed MAR mechanisms and MNAR mechanisms not only depending on the column itself can be considered. To translate **y** to a vector of missingness probabilities **p** ∈ [0, 1]^*n*^, we propose to use a 2-knot piecewise-generalized-sigmoid (pgs) function, cf. (8) further below. The first advantage of the 2-knot pgs function is that we only need the knot point and a point to the right and to the left of the knot point to define it. In fact, to make the construction scale-invariant the *x*-value of the points to the left and right are set to the minimum and maximum of **y**, leaving a total of 4 values for the user to pick. Two of these values, the corresponding *y*-values of the outer points are easy to interpret: they are the maximal and minimal probability for an entry to be missing in the column to be masked. The last two values – defining the knot point – are necessary to give the user the flexibility to decide how the slope should look. As seen in Figure 4 the above construction can be tuned from approximating a linear function, cf. Figure 4 B, to approximating a step function, cf. Figure 4 D. This leads us to the following definition of a *missingness scenario*.

**Figure 4:**
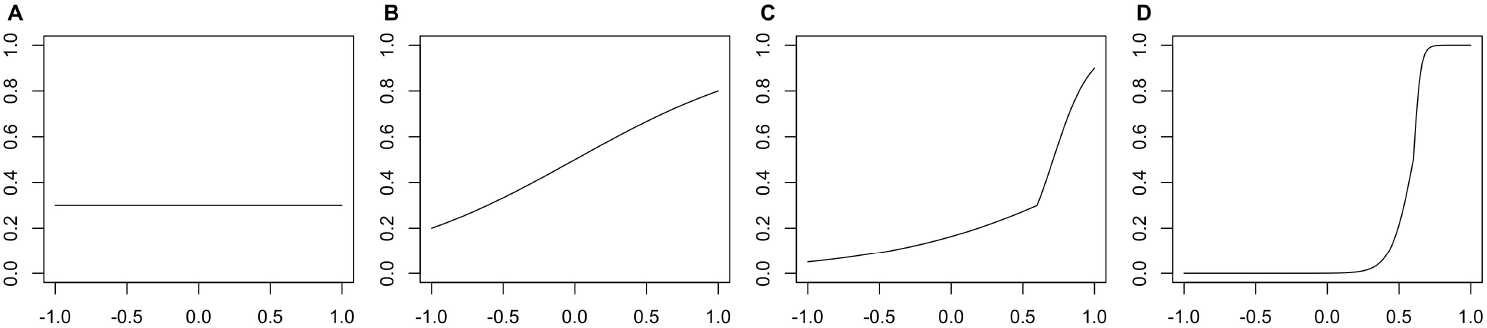
Examples of the 2-knot piecewise-generalized-sigmoid function 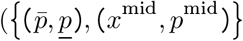 equal to {(0.3, 0.3), (0.5, 0.3)} (A), {(0.8, 0.2), (0.5, 0.5)} (B), {(0.9, 0.05), (0.8, 0.3)} (C), {(1 – 10 ^−7^, 10 ^−9^), (0.8, 0.5)} (D)), the x axis is scaled by the maximum value 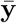 at 1 and the minimum value y at −1.

##### Definition 6.3.

Let **X** ∈ (ℝ ∪ {NA}) ^*n*×*p*^ be a data set, possibly with missing entries. A *missingness scenario* Ξ on **X** consists of

i. Three sets of indices ℐ_MCAR_, *ℐ*_MAR_, *ℐ*_MNAR_ ⊂ {1, 2, …, *p*} denoting the set of columns that should be masked via a MCAR, MAR and/or MNAR mechanism.
ii. For each *j* ∈ ℐ_*MCAR*_ a probability parameter *η*_*j*_ ∈ [0, 1], defining the average amount of MCAR missing entries.
iii. For each index *j* ∈ *ℐ*_MAR_ sets of indices 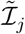 encoding the regressor-columns used to derive the missingness pattern in column *j*.
iv. For each index *j* ∈ *ℐ*_MAR_ a tuple 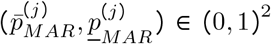 encoding the highest and the lowest probability of an entry being missing, as well as a Tuple 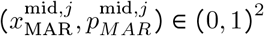 defining the knot of the pgs function.
v. For each index *j* ∈ ℐ_MNAR_ a tuple 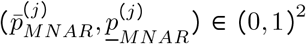 Encoding the highest and the lowest probability of an entry being missing, as well as a tuple 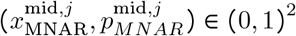 defining the knot of the pgs function.

Let us formalize the pgs function in the following, dropping the subscripts and index *j* in the following for the sake of readability. Let **y** be the sum of the standardized column vectors regressed upon (MAR) or the column-vector to be masked (MNAR), 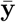 and **y** denote its maximum and minimum, respectively, and given the generalized sigmoid function by

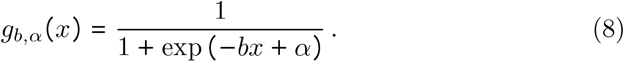

We compute a 2-knot psg function of the form (8) going through (**y**, *p)* and 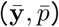 and with a knot at 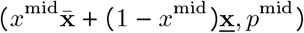 by

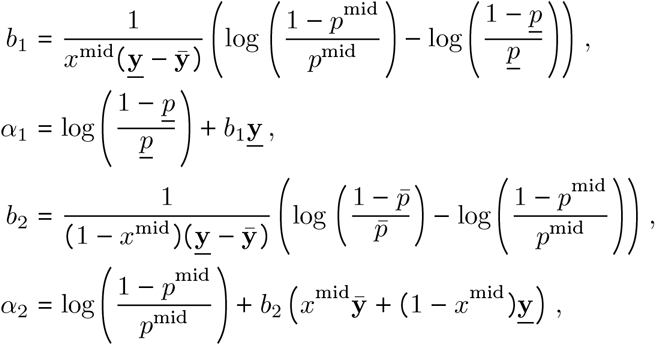

yielding

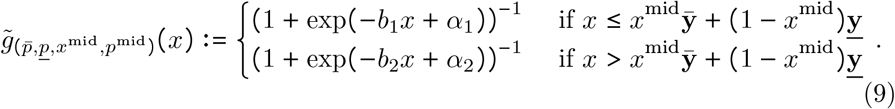

The vector of probabilities derives is computed by

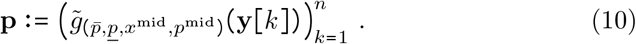

#### Missingness Pattern Simulation Protocol

Having defined a missingness scenario Ξ we give int he following the pseudo code of how to draw **M** from a data matrix **X** and Ξ.

First, initialize three matrices **M**_1_ and **M**_2_ and **M**_3_ as the one-matrix, i.e. **M**_*j*_[*k, ℓ*] = 1 for all *j, k, ℓ*. Moreover, denote in the following by 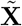 the column standardized matrix **X** (subtracting column-wise the sample mean and dividing by the sample standard deviation).

(1.) **For** *j* ∈ *ℐ*_MCAR_ **do:**

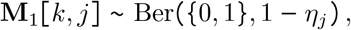

for all 1 ≤ *k* ≤ *n* independently.
(3.) **For** *j* ∈ ℐ_MAR_, **do:**
  a. Compute 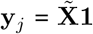 the vector of sums over the regressor columns.
  b. Compute the vector of success probabilities **p**_*j*_ ∈ 0, 1 ^*n*^ by (10).
  c. Draw

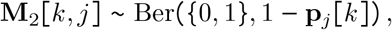

for all 1 ≤ *k* ≤ *n* independently.
(4.) **For** *j* ∈ *ℐ*_MNAR_, **do:**
  a. Set **y** := **x**_*j*_ where **x**_*j*_ is the *j*-th column of **X**
  b. Compute the vector of success probabilities **p**_*j*_ ∈ [0, 1] ^*n*^ by (10).
  c. Draw

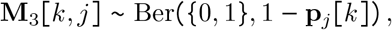

for all 1 ≤ *k* ≤ *n* independently.
(5.) Compute **M** as the entry-wise minimum of **M**_1_, **M**_2_ and **M**_3_.

#### Simulating a Missingness Scenario

It remains to detail how missingness scenarios can be drawn by more general *missingness parameters*. To this end, Table 4 defines the *missingness parameters*, which to a large extend are similar to the ones of a missingness scenario. The main difference of the parameters of Table 4 and Definition 6.3 is that columns to be masked and the columns to be used in the MAR regression are drawn at random from a set of columns. The remaining parameters defining the pgs functions are chosen once per mechanism instead of being chosen for each column independently. Hence, drawing a missingness scenario from missingness parameters is done in the following three steps.

**Table 4:**
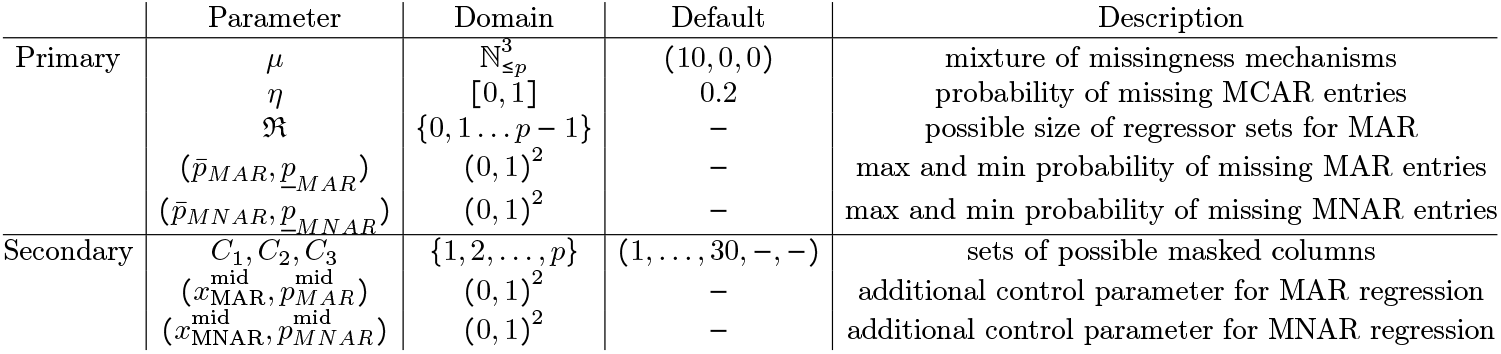
Missingness parameters *θ*_*M*_ used to simulate missingness scenarios.

(1.) The vector *µ* of Table 4 encodes via *µ*_1_, *µ*_2_ and *µ*_3_ the amount of columns featuring MCAR, MAR and MNAR missing entries, respectively. Via *C*_1_, *C*_2_ and *C*_3_ the user can restrict the choice of columns for each ingness mechanism. To construct *ℐ* _MCAR_ sample *µ*_1_ columns from *C*_1_ ⊂ miss- {1, 2, … *P* } without replacement. Analogously draw *ℐ*_MAR_ and *ℐ*_MNAR_ from *C*_2_ and *C*_3_, respectively.
(2.) Set *η*_*j*_ = *η* for all *j* ∈ *ℐ* _MCAR_ . Moreover, set 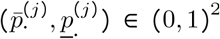 and 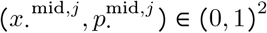 to be 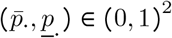 and 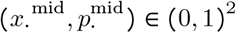 for both · = MAR and · = MNAR and for all *j*.
(3.) For each *j* ∈ *ℐ* _MAR_ draw *n*_MAR_ uniformly from ℜ ⊂ {0, 1, 2, …, *p* − 1}. Then draw *n*_MAR_ regressor columns sampled from {1, 2, …, *j* − 1, *j* + 1, …, *p*} without replacement to constitute 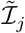.

### 6.3 Further Choices Made in ImputeBench

After having defined the data simulation and the missingness pattern simulation protocols used by the package `ImputeBench` the remaining inputs of the ImputeBench benchmarking protocol, cf. Figure 1, are largely provided by the user. To the end of making the package rather straight-forward to apply, however, there have been default imputation methods and their training functions included in `ImputeBench` and the choice of loss function ζ has been limited.

#### Default MVI Methods and their Training Functions

`ImputeBench` includes four default MVI-methods: k-nn impute, MICE, missForest and soft impute, for details see [86, 39], [88], [83] and [56] and Appendix A of the Supplementary Materials, as these are four of the most used and/or compared MVI methods, cf. Section 4, and their implementations are publicly available in the R-packages `VIM`, `mice`, `missForest` and `softImpute`, respectively. While for k-nn impute and soft impute there is one main parameter to be trained, *k* in the case of k-nn impute and *λ* in the case of soft impute, the customization of MICE and missForest is either not as straight-forward (MICE), or, not essential (miss-Forest). Thus, for now, `ImputeBench` trains *k* and *λ* of k-nn impute and soft impute, respectively, via a bisection algorithm and sets default values for MICE and missForest, details are given in Appendix C of the Supplementary Materials. Meanwhile, as baseline MVI method the user can choose between median and mean imputation.

##### Note 6.4.

*Within* `ImputeBench` *integer and binary columns are detected and all MVI methods - provided by the user or default - are rounded or binarized on the respective columns recognized as integer or binary*.

#### Loss Function

We propose to use a scaled root-mean-squared-error (RMSE) for `ImputeBench`. Given the data matrix **X** ^(*τ)*^ and an imputed data matrix **X** ^(*τ)*^, cf. Figure 1, compute first the vector of empirical standard deviations **v** ∈ ℝ^*p*^ of the columns of **X** ^(*τ)*^ . Second, let *ℳ* be the set of missing entries, compute

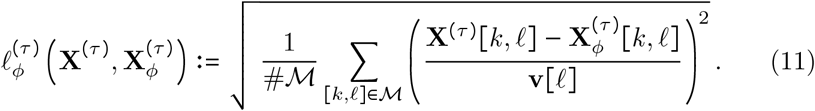

In the case **X** ^(*τ)*^ is simulated, additionally errors over all columns of the same “data type”, compare ν in Section 6.1, are provided by `ImputeBench`. For real data sets `ImputeBench` includes the additional option to partition the columns of **X** into *error groups*, these are mutually disjoint subsets *E*_1_, *E*_2_, …, *E*_*J*_ ⊂{1, 2, …, *p*} such that 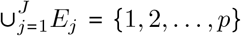.The error group losses 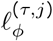 are computed via (11) for each group *j* = 1, 2, …, *J*, individually, and provided as an output of `ImputeBench`. Moreover, the overall loss is computed as the average over all error group losses by

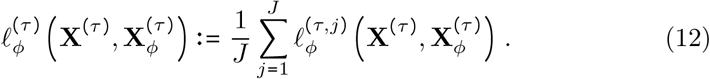

While the RMSE is a canonical choice as a loss, by normalization with **v** in (11) the loss becomes scale-invariant and each column is given the same weight in the overall loss. Moreover, via the introduction of error groups in its grouped version in (12) the user can consider columns singularly and weight them in the reported overall loss. Further popular losses, such as an *ℓ*_1_-type loss, might be added in future versions of the `ImputeBench` package. Moreover, note, as later discussed in Section 7, assessing MVI methods performance just reduced to a single loss value might be too reductive at times, then visual inspection of imputed values can shed more light on the performance of an MVI method.

## 7 Simulation Study

We exemplify in this section the use of `ImputeBench` on three simulated data examples and one real world data example, given by data from a food frequency questionnaire (FFQ). The FFQ data is used to suggest a work-flow on how to use `ImputeBench` in practice.

### 7.1 Simulated Data

As emphasized throughout this paper the amount of data-missingness-distributions is vast and thus the three given simulated imputation benchmarking examples given in Figures 5, 6 and 7 can only showcase selected, interesting behaviours of MVI methods. The parameter choices for the three settings are given in Table 5. Using these examples we will make in the following some observations, regarding the package `ImputeBench`, which implements the benchmarking protocol of Section 5 with the data and missingness pattern simulation protocols of Section 6, and, MVI benchmarking in general.

**Table 5:** Parameters of the examples in Figure 5, 6 and 7. An *x* stands for a parameter that is varied in its respective figure. Non-mentioned parameters are said to their respective default values reported in Table 3 and 4.

| Parameter | Figure 5 | Figure 6 | Figure 7 |
| --- | --- | --- | --- |
| $n$ | 500 | $x$ | 500 |
| $p$ | 100 | 50 | 100 |
| $A$ | 1 | 1 | 2 |
| $q$ | 10 | 20 | $x$ |
| $U$ | 1 | 1 | 1 |
| $(u_1, u_2)$ | (0.5, 1.5) | (0.1, 1.5) | (0.1, 1.5) |
| $\sigma$ | 0.25 | 0.25 | 0.5 |
| $\rho$ | 0.4 | 0.4 | 0.4 |
| $\alpha$ | 0 | 0.3 | 1 |
| $\nu$ | (100, 0, 0, 0, 0, 0) | (50, 0, 0, 0, 0, 0) | (0, 50, 50, 0, 0, 0) |
| $\mu$ | (0, 0, $x$) | (50, 0, 0) | (0, 0, 100) |
| $C_1, C_2, C_3$ | $\{1, \dots, x\}$ | $\{1, \dots, 50\}$ | $\{1, \dots, 100\}$ |
| $\eta$ | — | 0.2 | — |
| $(\bar{p}_{MNAR}, \underline{p}_{MNAR})$ | (0.9, 0.1) | — | (0.8, 0.05) |
| $(x_{MNAR}^{\text{mid}}, p_{MNAR}^{\text{mid}})$ | $(0.5, 0.4)^2$ | — | (0.7, 0.3) |

**Figure 5:**
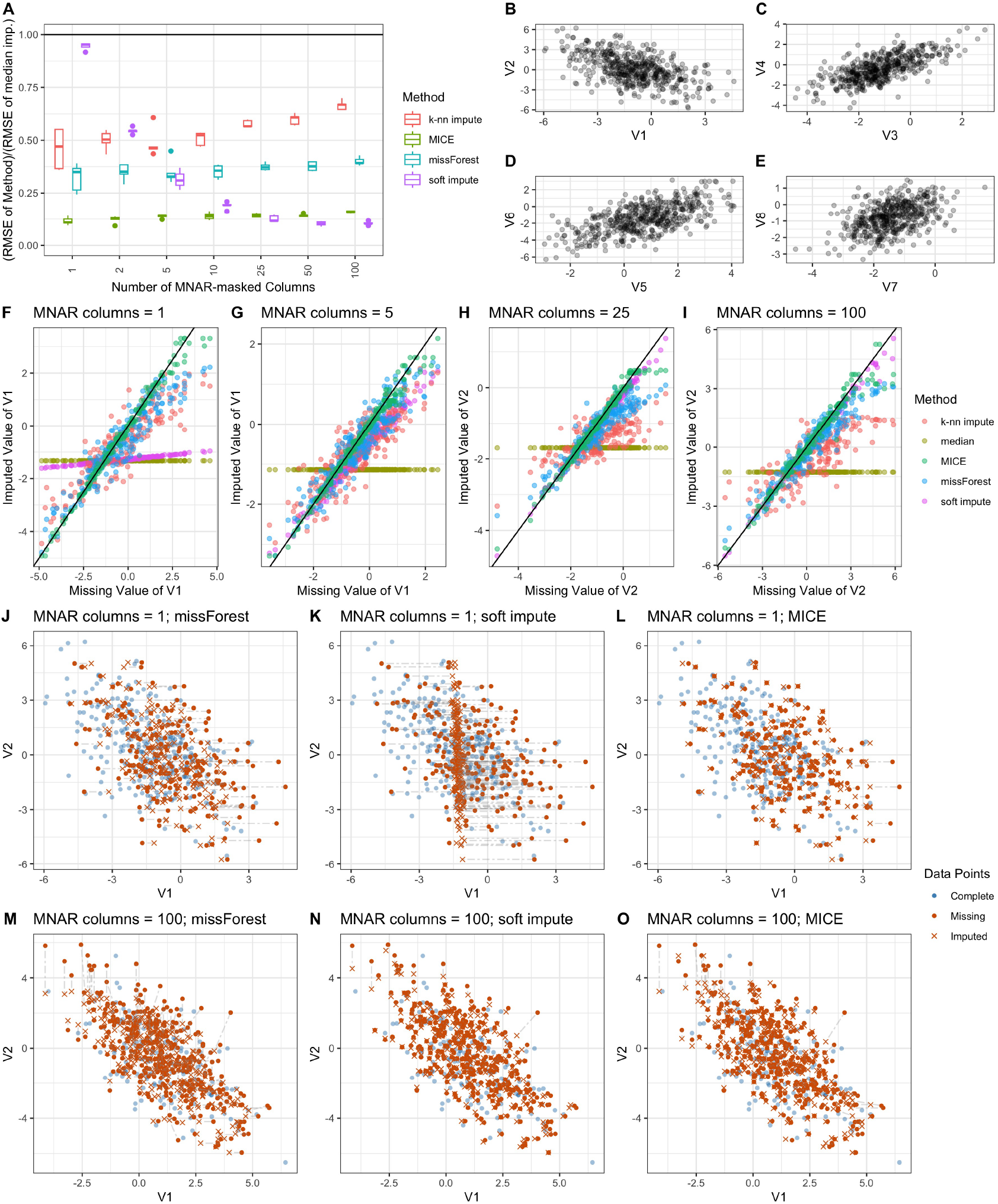
Imputation performance of k-nn impute, MICE, missForest and soft impute under parameter choices reported in Table 5, varying the number of MNAR masked columns (A,F-O). Examples of the drawn data (B-E)

**Figure 6:**
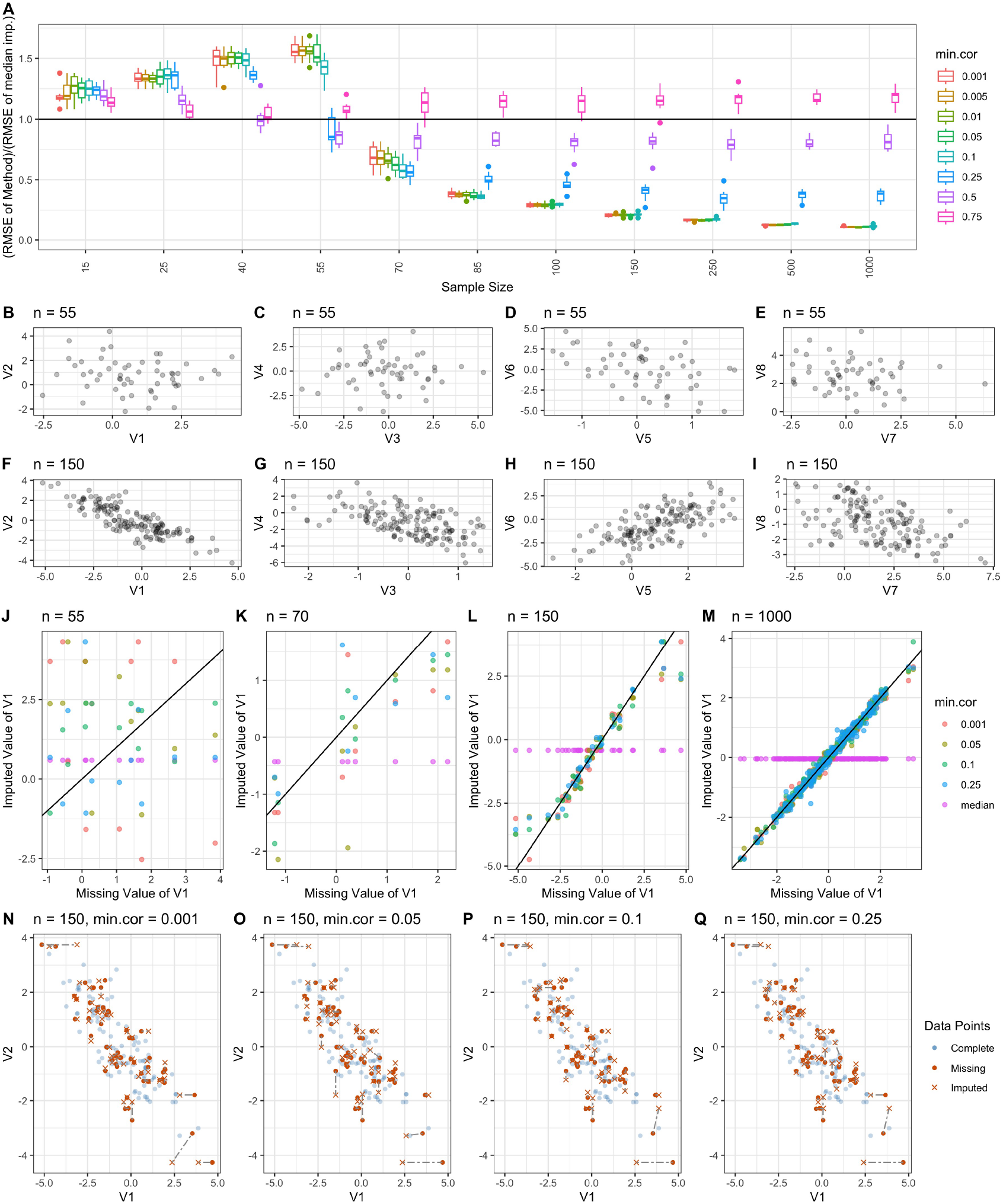
Imputation performance of MICE for different values of `min.cor` under parameter choices reported in Table 5, varying the sample size *n*(A,J-Q). Examples of the drawn data (B-I).

**Figure 7:**
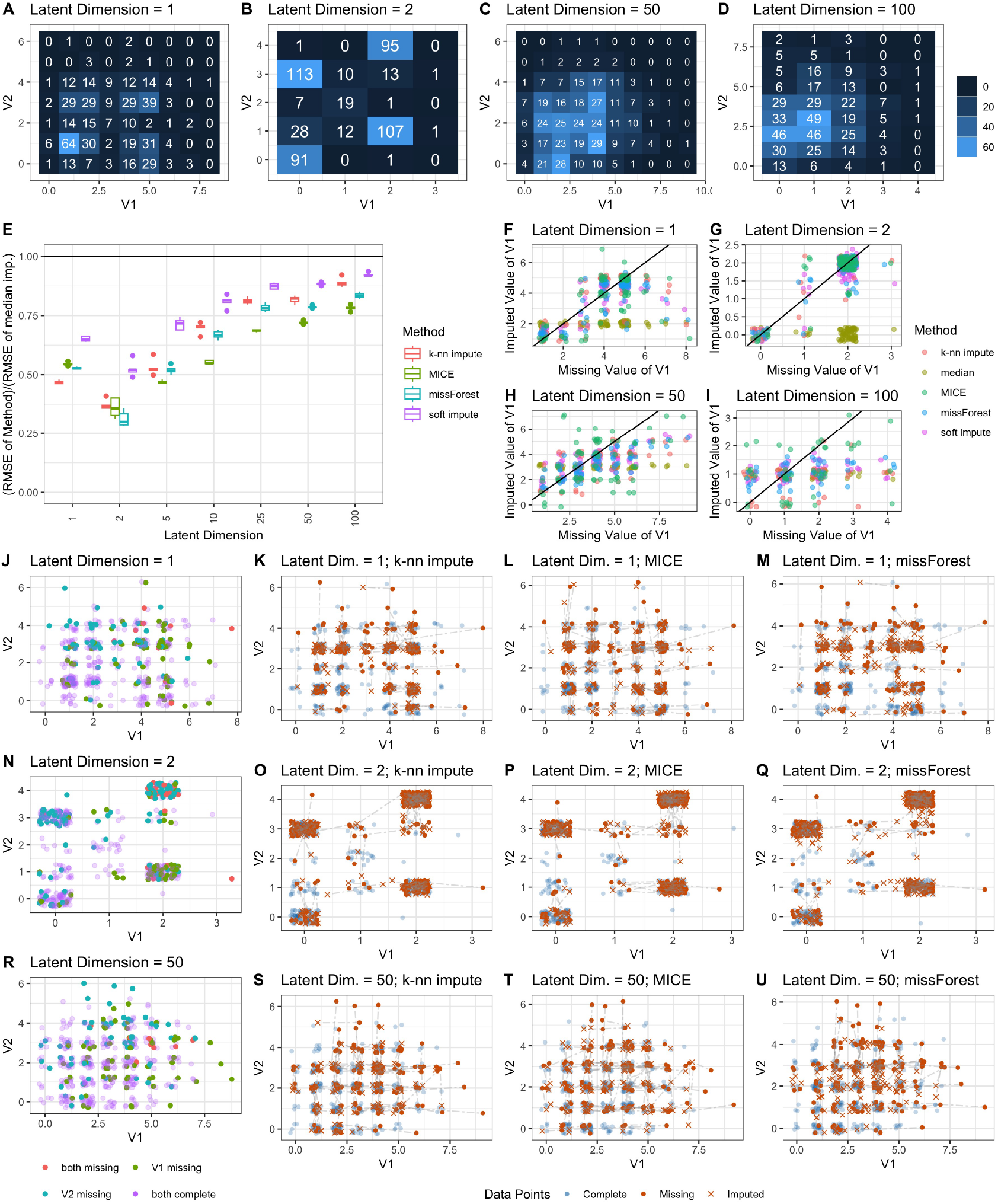
Imputation performance of k-nn impute, MICE, missForest and soft impute under parameter choices reported in Table 5, varying the number of MNAR masked columns (E-I,K-M,O-Q,S-U). Examples of the drawn data (A-D) and of the drawn missingness pattern (J,N,R).

#### ImputeBench Allows for a Variety of Data Distributions

As examples of the variability of `ImputeBench` see in Figure 5 B-E draws of a “standard” normal distributed model with some low-dimensionality and no featured non-linearity and in Figure 7 A-D draws of a Poisson/Bernoulli distributed model with a spherical shape (*α* = 1) and varying low-dimensionality. As discussed in Section 6.1, more data features such as multi-modal and heavy-tailed marginal distributions and spline- and sine-transformed variables can be used to customize the data simulation protocol, cf. Figure 2. Since the design of “believable” simulated data is hard, especially in a high dimensional setting (*p* large) where visual inspection is only available for the marginal distributions, we argue that including as many tuning parameters as `ImputeBench` does, cf. Table 3, is paramount to be sufficiently fine-tunable.

#### Different Data-Missingness-Distributions Call for Different MVI-Methods

Compared in Figure 5 and 7 are the four default MVI-methods included in ImputeBench: k-nn impute, MICE, missForest and soft impute. Recall that only the *k* of k-nn impute and the *λ* of soft impute are trained in these examples, cf. Section 6.3. Hence, the following remark.

##### Remark 7.1.

*The given examples of Figure 5, 6 and 7 are <u>not an exhaustive</u> benchmarking of the four default MVI methods k-nn impute, MICE, missForest and soft impute on the given simulated data, as in particular the latter two are not optimized for performance*.

However, even without elaborate fine-tuning it can be said that for each of the four default MVI methods a data-missingness-distribution can be found in Figure 5 A and Figure 7 E under which each respective MVI method outperforms the others. This reaffirms the mixed results in the surveyed benchmarking papers in Tables 1 and 2 of Section 4.

#### ImputeBench can be Used to Optimize MVI Methods

In Figure 6 MICE performance is compared for different choices of its parameter `min.cor`, which in effect controls the minimal correlation with the target variable a column must have to be included in the regression model, for details on `min.cor` we refer to [89]. As sample size is increasing, the underlying latent model becomes more and more apparent when visualizing the data (Figure 6 B-I) and around *n* ∼ 55, 70 MICE starts to outperform median imputation depending on the choice of `min.cor`. Moreover, the optimal choice of `min.cor` seems to also depend on the sample size, for example `min`.`cor` = 0.25 performs best for *n* = 70, but from *n* = 85 onwards it performs considerably worse than lower choices of `min.cor`. Considering, the plots of missing vs. imputed values in Figure 6 J-M, one notices that for higher sample sizes while higher values of `min.cor` also pick up the latent low-dimensional subspace the data lies on they do so with an increased variance of the error.

#### Visualizing the Imputed Values Is Important

In Figure 5A we observe a seemingly counter-intuitive result: the performance of soft impute in relation to median imputation *improves* as more columns have missing entries, i.e. as more entries are missing in the data matrix. Such an example shows that closer inspection of the imputed values as given in Figure 5 F-O is often necessary, if only as a sanity check, as only considering the performance measure might only reveal a part of the story. In this case, having only one column with MNAR-missing entries instead of all hundred columns seemingly harms soft impute, as it tries to fit - in its default version - a matrix with low nuclear-norm by an objective function that equally weights the error on entries from columns without missing entries and the error on entries from (the) column(s) with missing entries, see [56] for details. As by construction (the) column(s) with missing entries has/have fewer observed entries, its(their) importance in the objective function is down-weighted. Consequently the variance of (the) column(s) with missing entries is(are) largely ignored and the imputed values are close to the median imputation, cf. Figure 5 F,K. This effect disappears as missingness is more evenly distributed over all columns.

Moreover, in particular in the MAR and MNAR setting visual inspection of the imputed values can add to the understanding how MVI methods perform when they have to extrapolate, i.e. when tail values of the distribution are missing. For example in Figure 5 O, Figure 6 P,Q and in Figure 7 T one can see that MICE seems to be better at extrapolation, as missing tail values are not necessarily pushed towards the median, but might even be imputed a bit further out.

### 7.2 Real Data

To find an appropriate MVI method for a given data set via `ImputeBench` we propose in the following a potential work-flow and exemplify this work-flow with a real life example. Note that, as every data set and task underlies different cost and time constraints the work-flow proposed below can only serve as a guide.

#### Work-Flow

##### 1 Design plausible missingness scenarios

Deriving appropriate missingness mechanisms can be done either by using statistical test on MAR missingness, see for example [14, 67, 47], or by less time-consuming visual inspection of the data and the missingness pattern. Note that as the actual missing values are unknown there is no possible test for MNAR missingness, in this case one needs to resort to visual inspection and external assumptions. Visual inspection can be performed by plotting the missing pattern of a particular column against the observed values of different columns (see Figure 8 B-D).

**Figure 8:**
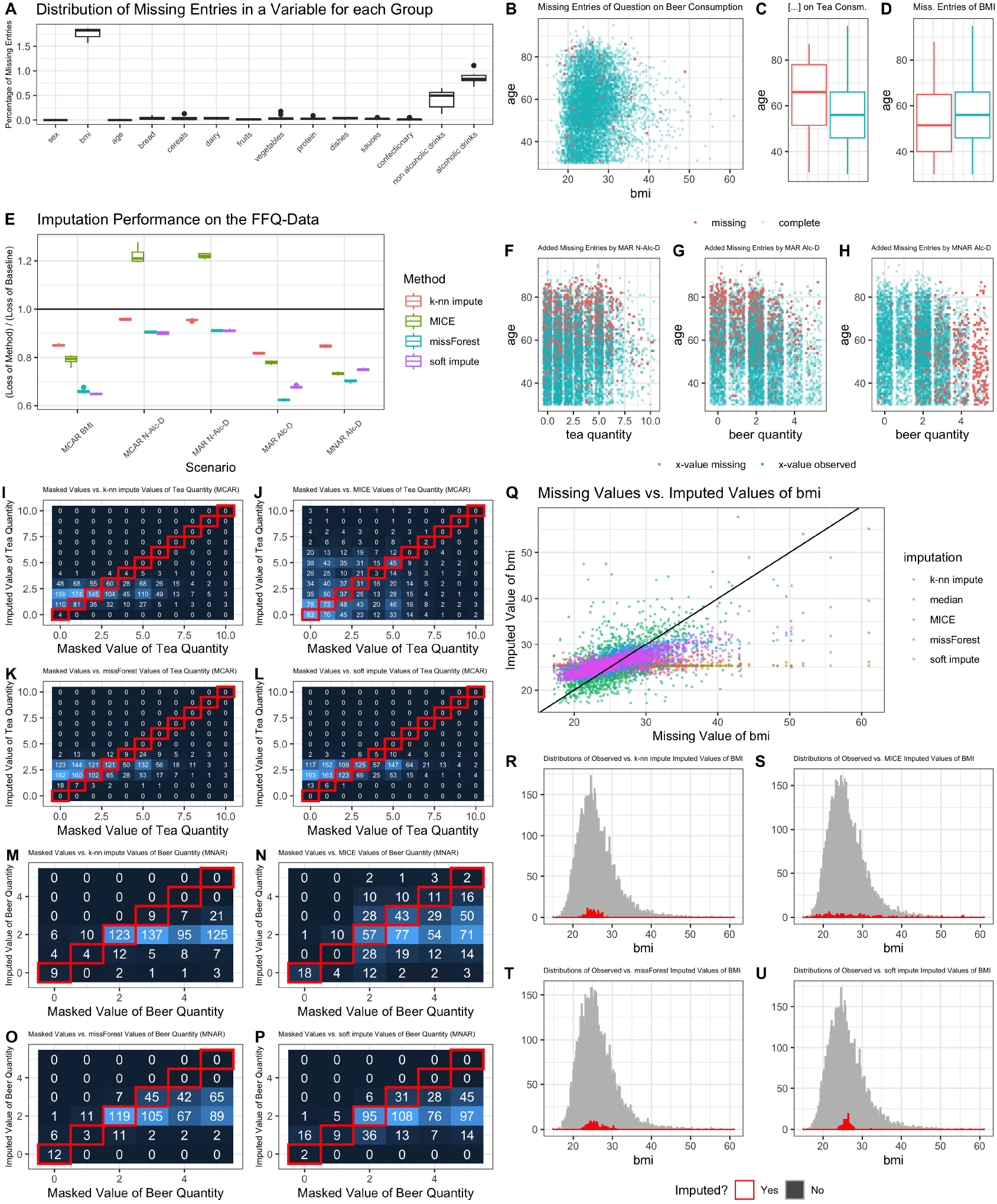
Imputation performance on the FFQ-data under missingness scenarios reported in Table 6 (E). Distribution of missing entries (A); real missingness pattern (B-D); checking the missingness scenarios (F-H); visual checks of the imputed values (I-U).

##### 2 Test the missingness scenarios

We recommend, in particular when designing MAR and MNAR missingness scenarios, to create test masks and plotting their distribution against the non-masked values (see Figure 8 F-H). The choices of 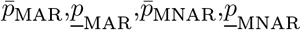 and 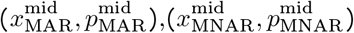 and their actual impact on the drawn missingness pattern are far from being transparent in particular with high-dimensional data, cf. Defini-tion 6.3. Hence, visual inspection helps to fine-tune these parameters.

**Table 6:**
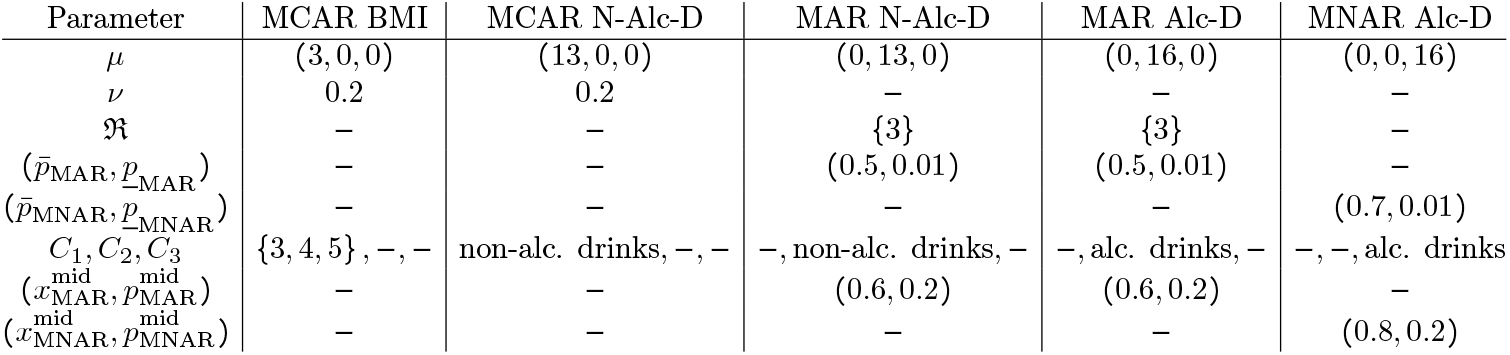
Missingness scenarios chosen for the FFQ-Data.

##### 3 Choose suitable MVI methods and their parameter training functions

`ImputeBench` usesmedian imputation as the baseline MVI method and four further default methods: k-nn impute, MICE, missForest and soft impute, see Section 6.3 for details. However, other approaches or other configurations of the default MVI method might be appropriate for the data distribution at hand and should be taken in consideration. Customized MVI methods can be passed to `ImputeBench`, we refer the reader to the accompanying vignette of `ImputeBench`.

##### 4 Choose a suitable loss function

For now `ImputeBench` supports only a standardized RMSE as a loss function, which is customizable only via grouping columns into *error groups*, see Section 6.3 for details. We recommend considering whether the RMSE fits the given task and to consider computing the loss also on single variables/columns, again we refer to the vignette of `ImputeBench` for details.

##### 5 Find the MVI method that performs best

The `ImputeBench` package outputs an overall performance over the whole matrix (usually the average over all error groups) and a performance on each error group (see Figure 8 E). In the best case one MVI method outperforms on all scenarios (and error groups). Otherwise, diverging performance on different variables (or groups) or scenarios can indicate that a combination of MVI methods might be beneficial, which could be evaluated in another run of `ImputeBench`.

##### 6 Visual checks

Once an MVI method is chosen in Step 5., we recommend two visual checks of the imputed values. The first is given by considering the imputation performance on the additionally drawn missingness patterns governed by the chosen missingness scenarios (see Figure 8 I-Q). The second is given by plotting the distribution of imputed values (on “real” missing values) against the distribution of the observed values (see Figure 8 R-U). While the first check gives a broader insight in how the imputed values behave in particular in terms of bias-variance-trade-off, the second check visualizes how the imputed values fit to the distribution of the observed values.

#### Example - FFQ Data

To exemplify the above work-flow we apply `ImputeBench` in Figure 8 to answers of a food frequency questionnaire (FFQ) undertaken in the Rhineland Study, see [7], a large-scale population-level study in Bonn, Germany. The FFQ data consists of 7842 rows of individual participants and 141 columns, 5 of which correspond to *age, bmi, height, weight* and *sex* of the participants and 136 of which correspond to multiple-choice responses to the FFQ. Subjects with missing entries exceeding a threshold of 20% had been excluded from the data set in a pre-processing step, the remaining data consists of very few remaining missing entries, cf. Figure 8 A, that are to be imputed.

Applying the work-flow from above to the data set we first design missing scenarios by considering in which food groups considerable amount of missing values appear (Figure 8 A) and by visual inspection as in Figure 8 B-D. To this end, we design 5 scenarios formalized in Table 6: two MCAR mechanism on *bmi* (MCAR BMI) and on questions relating to the food group of *non-alcoholic drinks* (MCAR N-Alc-D), two MAR mechanism on questions relating to the food group of *non-alcoholic drinks* (MAR N-Alc-D) and *alcoholic drinks* (MAR Alc-D) regressed on *age* and an MNAR mechanism on the food group of *alcoholic drinks* (MNAR Alc-D). Scenarios (MCAR BMI) and (MCAR N-Alc-D) do not make any specific assumption on the missingness pattern. Meanwhile, Scenarios (MAR N-Alc-D) and (MAR Alc-D) are derived from visual inspections such as depicted in Figure 8 C showing the age distribution for existent and missing values in the *quantity of tea* variable, and, the external assumption that the memory capacity of humans decreases with age. Last, Scenario (MNAR Alc-D) is reflecting the external assumption that there might be a social stigma related to the high consumption on alcohol and the observation that missingness is most prevalent in the questions on *alcoholic drinks*, see Figure 8 A.

In Figure 8 F-H tests of the MAR and MNAR missingness scenarios are displayed using the variables *tea quantity* and *beer quantity*. The fine tuning was done so that we have a visible gradient between *age* or *beer quantity* and missingness in the target variable, respectively, while not masking all extreme cases (in particular in Figure 8 H).

Being an example of the work-flow the performance comparison in Figure 8 E on the FFQ-data cannot be considered exhaustive, as only default MVI methods are considered (often using default values of tuning parameters). In particular, the relatively poor performance of MICE on the *non-alcoholic drinks* group suggests that further tuning of the MICE parameters would be useful. Meanwhile, for the loss function, the data columns were grouped into respective food groups (whilst keeping *age, sex* as single column error groups and grouping together *bmi, weight* and *height*), cf. Figure 8 A.

Overall we see that missForest shows promising results with regard to the standardized RMSE loss, see Figure 8 E. Considering the visual checks one can see in Figure 8 Q very well that while MICE is good at even recovering high values its error stems largely from a high variance, meanwhile in the order of soft impute, missForest and k-nn the introduced bias increases and large values can not be recovered. The same is true to a different extent for the discrete variables depicted in Figure 8 I-P. As MICE performs similarly strong as missForest and soft-impute in Scenario (MNAR Alc-D) one could argue in favour of MICE as its variance seems to model best the true underlying variance (see Figure 8 N vs. Figure 8 O and P). Last by comparing the observed and imputed value distributions of the *bmi* variable in Figure 8 R-U one finds the distribution of the MICE imputed values of *bmi* to be furthest away from the distribution of the observed values.

Last, we make a recommendation regarding the computational costs.

##### Note 7.2.

*In case the computation time of* `ImputeBench` *is not acceptable one might consider excluding default MVI methods, replacing the training functions by simpler ones, reducing the number of runs T, or, sub-sampling the rows of the given data matrix. Otherwise, both on simulated and on real data the benchmarking functions of* `ImputeBench` *allow to specify a maximal run-time per MVI method*.

## 8 Conclusion

Benchmarking missing value imputation methods is a not straight-forward endeavour and should be considered on a case-by-case basis. To help make benchmarking of MVI methods on a case-by-case basis feasible we proposed in this paper a protocol on benchmarking MVI methods called ImputeBench, cf. Section 5. We then went on to propose a specific implementation of the ImputeBench protocol as the `ImputeBench` R package, this included the definition/development of

- a data simulation protocol;
- a way to draw missingness patterns via the notion of a *missingness scenario* and a missingness pattern simulation protocol;
- a missingness scenario simulation protocol;
- a general loss function.

In the interests of clarity and well-definedness, we revisited the definitions of missingness mechanisms in Section 3 and proposed a slight refinement of these by *column MAR* and *column MNAR*. Last, in Section 7.2 a work-flow to practitioners on how to use `ImputeBench` was given.

Benchmarking of MVI methods cannot be entirely exhaustive and similarly no data and missingness pattern simulation protocol employed for the ImputeBench benchmarking protocol can be exhaustive either. Nevertheless, by defining a protocol such as ImputeBench and developing its accompanying implementation `ImputeBench` our intention is to make benchmarking of MVI methods easier, more transparent, reproducible and as broad as practically possible. Moreover, we hope that for the development of future MVI methods `ImputeBench` can be employed to test their performance in a controlled setting, including regimes that challenge existing and emerging MVI methods.

We mention in conclusion two specific future directions that would be needed to address specific limitations of this work. First, at the moment we do not ask whether CCA might be an option to answer the question *“Which entries to impute?”* Second, the MVI methods designed for multiple imputation have to be used as single imputation methods. To extend `ImputeBench` to include CCA and multiple imputation methods the main requirement would be modification of the loss function defined in (11) and (12), as performance would need to be measured indirectly via a subsequent estimation or prediction task. Hence, assumptions on the later use for estimation/prediction would need to be provided.

## Data Availability

The accompanying R package ImputeBench is available on github: https://github.com/richterrob/ImputeBench
and its vignette is available on figshare: https://figshare.com/articles/online_resource/ImputeBench_Vignette_html/23896677.
The data from the Rhineland Study used in this manuscript is not publicly available due to data protection regulations. Access to data can be provided to scientists in accordance with the Rhineland Study's Data Use and Access Policy. Requests for additional information and/or access to the datasets of the Rhineland study can be send to.

https://figshare.com/articles/online_resource/ImputeBench_Vignette_html/23896677

## 9 Acknowledgements

This work was supported in part by the German Federal Ministry of Education and Research (BMBF) project “MechML” and the Diet-Body-Brain Competence Cluster in Nutrition Research funded by the German Federal Ministry of Education and Research [‘01EA1410C’,’01EA1809C’ to M.M.B.B.)

## 10 Supplements - Package and Vignette

The accompanying R package `ImputeBench` is available on github: https://github.com/richterrob/ImputeBench and its vignette is available on figshare: https://figshare.com/articles/online_resource/ImputeBench_Vignette_html/23896677.

## A Overview of MVI Methods

In Tables 7 and 8 an overview of the main MVI methods considered in the benchmarking papers of Section 4 can be found. Note, that the given overview is broad but can not be comprehensive due to the vast amount of papers on MVI. It aims at clustering methods in rather rough categories in order to ease the comparison of benchmarking papers in Section 4. For a recent more detailed review of MVI methods we refer the reader to [50]. As it would exceed the scope of this paper to go into the technical details of the MVI methods referenced, in the remainder of this appendix only we give a high level description of the different MVI method cluster.

**Table 7:**
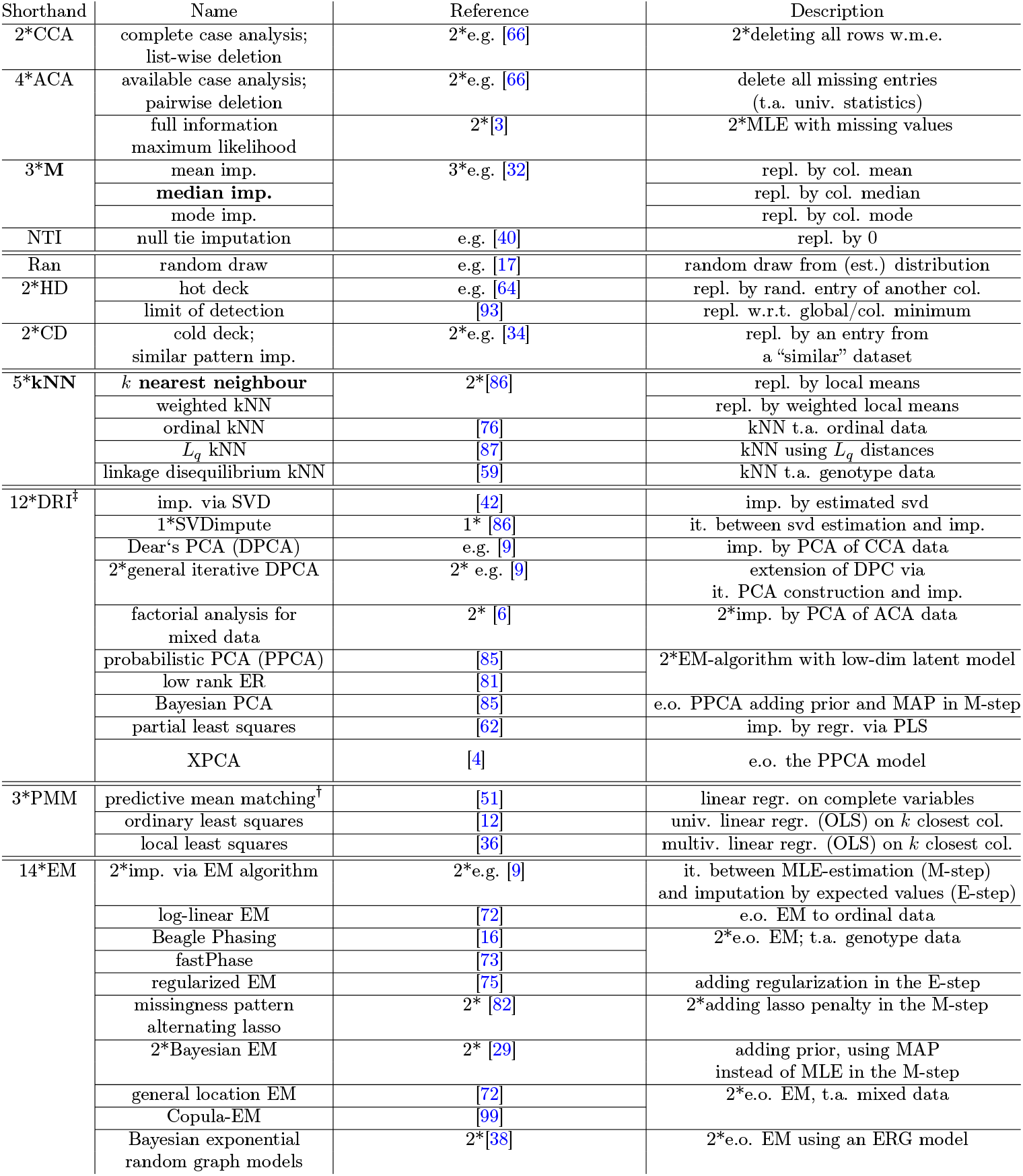
MVI methods († indicates developed as a multiple MVI, **bold methods** are included as defaults in ImputeBench. Abbreviations used: imp. = *imputation*, e.o. = *extension of*, t.a. = *targeted at*, it. = *iterates*, w.m.e. = *with missing entries*, univ. = *univariate*, multiv. = *multivariate*, est. = *estimated*, rand. = *random*, repl. = *replacement*, w.r.t. = *with respect to*, regr. = *regression*, col. = *column(s)*, ‡DRI = *dimension reduction imputation*, ‡DLI = *deep learning imputation*.)

**Table 8:**
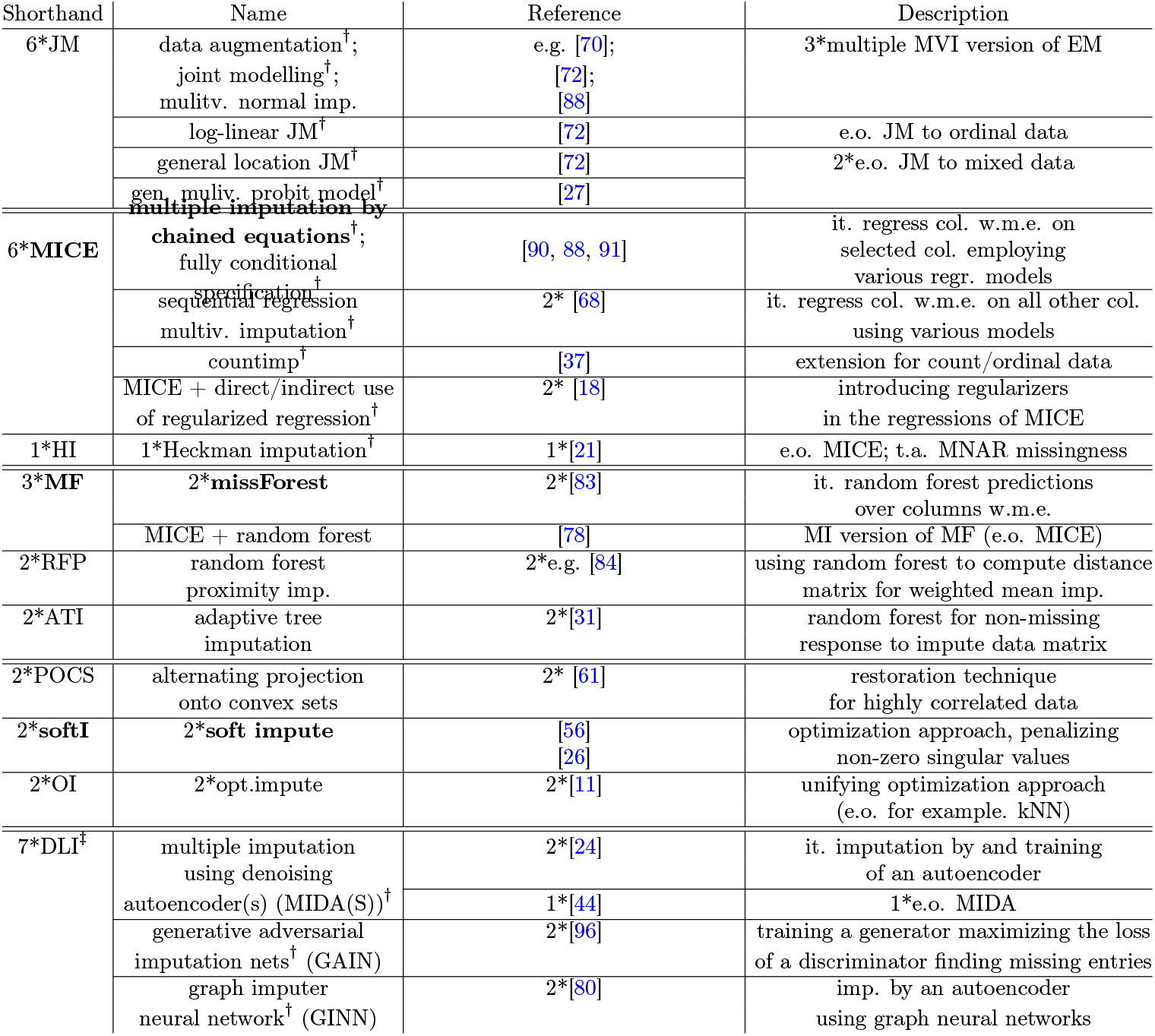
Continuation of Table 7.

### Exclusion/Ad hoc imputation (CCA, ACA, M, NTI)

The two main *ad hoc* approaches to deal with missing values are exclusion of all rows or entries with missing values as done in *complete* and *available case analysis*, and, imputation with a straight-forward statistic of the observed values of each column such as in *mean, median* and *mode imputation*. All of these approaches are conceptually straightforward, but suffer if the observed data is biased especially if the MCAR assumption is violated. Moreover, the mean, median and mode imputation have the disadvantage of underestimating the sample variance. A special case is the *null tie imputation* as it imputes each missing entry with 0, which is often the default method in experiments suffering from a limit of detection.

### Imputation by random draws (Ran, HD, CD

Imputation by random draws has a long tradition, here missing entries are drawn from a “suitable” predefined distribution, or, as in *hot* and *cold deck* methods from an empirical distribution chosen from the available data or from additional data. While such methods generally solve the problem of underestimating the variance present in mean/median/mode imputation, they often suffer from a lack of justification and performance in terms of imputation error, as well as, a lack of generality.

### Nearest neighbour imputation (kNN

This category comprises imputing missing entries by, possibly weighted, local means of *k* “neighbouring” data rows. *k*-nearest neighbour methods might vary concerning the distance measures employed to calculate the closest neighbours or concerning the introductions of weights. For a comparison of nearest neighbour imputation in practice see [10].

### Imputation by dimension reduction models (DRI

Under this cluster MVI methods are grouped that first estimate a latent model of reduced dimension from the observed data and then reconstruct the missing entries via the estimated latent model. In most cases the latent model is first estimated by a pre-imputed dataset, e.g. via some ad hoc MVI method, and then the method iterates between imputing the missing entries via the current latent model, and estimating the latent model with the current imputed values. Note, dimension reduction models for imputation are often used for multiple imputation. Among the most frequent dimension-reduction models used in MVI are the principal component analysis, singular value decomposition and partial least squares.

### Imputation by modelling the joint distribution (EM, JM

This group of MVI methods are largely based on imputing missing values by modelling the (joint) distribution of **X**_lat_ ∼ *𝒟*_lat_ - where *𝒟*_lat_ is parametrized by some *θ* - estimating its parameter(s) *θ* and imputing either by the derived expected value of the missing entries (single MVI), or, by drawing from the derived distribution (multiple MVI). To the end of single MVI, an *expectation-maximization* (EM) algorithm is usually employed, iterating between estimating the imputed values given the current *θ* (E-step) and maximizing an MLE- or MAP-estimator of *θ* given the current imputed values (M-step). The multiple MVI version of the EM algorithm also known as *joint modelling* or *data augmentation* yields an MCMC algorithm. To this end, a prior on *θ* needs to be specified beforehand. In the MCMC algorithm the E-step of the EM algorithm is replaced by drawing from the distribution *𝒟*_lat_ for the current *θ* and the M-step is replaced by drawing parameters *θ* from the posterior distribution on *θ* given the current imputed values. As the success of the imputation method now depends on the fit of the chosen parametric model of *𝒟*_lat_ to the data, there exists a multitude of joint modelling approaches proposed in the literature, in particular, aimed at categorical and mixed data. A caveat of modelling and estimating in each iteration the complete distribution *𝒟*_lat_ is the high computation complexity as the number of columns/features *p* of **X**_lat_ grows.

### Imputation as a parametric prediction problem (PMM, MICE, HI)

Parallel to the EM/JM approaches imputing missing values by regressing on all, or most often, a subset of “neighbour” columns, for example by local least squares, became popular. This cluster can be seen as a computationally more efficient counterpart of the above EM/JM approaches. Also for parametric regression approaches the fit of the model class to the data is key for its success as an MVI method. Meanwhile, multiple MVI via parametric regression methods is commonly performed by putting a prior on the regression coefficients and then drawing the imputed values as well as the regression coefficients of their respective distributions. Arguably the most popular representative of this cluster is *multiple imputation by chained equations* (MICE) also known as *fully conditional specification* which iterates over all column with missing entries and imputes via various user-specified regression models.

### Imputation as a non-parametric prediction problem (MF, RFP, ATI, DLI)

This cluster contains MVI methods based on non-parametric regression or machine learning techniques. In particular due to the increase in computational power these methods become more and more popular in recent years. One of the most successful methods of this cluster is *missForest*, which employs random forests for MVI. A main advantage of non-parametric methods is the lowered reliance on the correct specification of the regression/distributional models by the user, achieving often already good performance with default values. To the same end, in recent years deep learning architectures have been proposed for MVI, taking for example the form of autoencoders or generative adversarial nets. As with many ML approaches the main drawback is a certain in-transparency of the result, since reverse engineering which columns contributed in which way to the given imputation result is not straight-forward.

### Imputation by global constraints (POCS, softI, OI)

Taking ideas from signal processing, this cluster is composed of MVI methods building on optimization approaches. MVI methods in this cluster either assume highly correlated data (compare for example inpainting in image processing) or general global constraint on the latent dimensionality of the data. The latter is most prominently implemented in *soft impute*, an MVI method that minimizes an objective function balancing between a data fidelity term concerning the fit to the observed entries and an *ℓ*_1_-norm on the singular values of the data matrix (regularization term). Moreover, with *opt*.*impute* there exists a try to unify approaches in an optimization framework, including nearest neighbour and random forests imputation.

## B Transformation Stage Details on (2.)

In the following we give details on the exact (quantile-)transformations performed in step (**2**.) of the transformation stage of the data simulation protocol in Section 6.1. In particular, we discuss how exactly all secondary parameters of Table 3 of the main text come into play in the simulation protocol.

a. **Gaussian columns:** The first ν_1_ columns are quantile-transformed into Gaussian variables with random means drawn from *𝒩*(*g*_1_, *g*_2_) and random standard deviations drawn from *𝒩*_trunc_((0.5, ∞), *g*_3_, *g*_4_).
b. **Poisson columns:** The next ν_2_ columns are quantile-transformed into Poisson variables with random means drawn from Poi (*λ)* .
c. **Bernoulli columns:** The next ν_3_ columns are quantile-transformed into Bernoulli variables with random success probabilities drawn from *𝒰* ([*b*_1_, *b*_2_]), where *𝒰* denotes the uniform distribution.
d. **t-distributed columns:** The next ν_4_ columns are quantile-transformed into t-distributed variables with random degrees of freedom drawn from *𝒰 ({ t*_3_, …, *t*_4_}) and random means drawn from *𝒩* (*t*_1_, *t*_2_).
e. **Sine-transformed columns:** The next ν_5_ columns are sine-transformed by

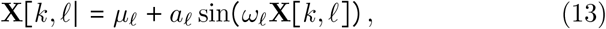

with *µ*_*ℓ*_’s drawn from 𝒩 (*ξ* _1_, *ξ* _2_), *a*_*ℓ*_’s drawn from 𝒰 ([*ξ* _3_, *ξ* _4_]) and *ω*_*ℓ*_’s drawn from *𝒩*._trunc_ ((ξ_5_,ξ _6_),ξ _7_,ξ _8_)
f. **Spline-transformed columns:** The next ν_6_ columns are spline-transformed by

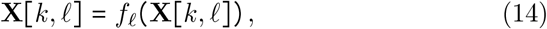

where the *f*_*ℓ*_’s are degree 1-splines (piecewise-linear functions) with *K* + 1 knots. Let *m*_1_ and *m*_2_ be the minimal and maximal value of **X** [*k*, ·], respectively, then the knots are given by

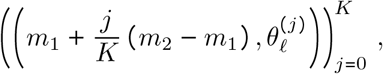

where the 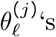 are drawn from

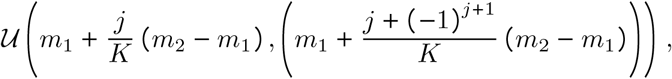

for all *j* and *ℓ*..
g. **The translation group column:** The last column is quantile-transformed into a discrete uniform distribution on {1, …, *U*} .

## C Details on the Training Functions Included in ImputeBench

In this appendix we detail in brevity the default parameter choices made by `ImputeBench` for the default MVI methods MICE and missForest, as well as, the training algorithms of the default MVI methods k-nearest neighbour impute and soft impute included in `ImputeBench`.

### C.1 k-nearest neighbour imputation

To include k-nearest neighbour imputation as a default MVI method in `ImputeBench` we use the R-package `VIM`, see [39]. All parameters but *k* are set to default values of `VIM::kNN`, the training of the parameter *k* by a bisection algorithm is detailed in the following.

Given a data matrix **X**_obs_ ∈(ℝ {*NA*}) ^*n*×*p*^ with missing entries. To train *k* of k-nearest neighbour imputation we start by defining the following parameters

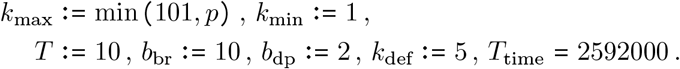

We define a default missingness parameters

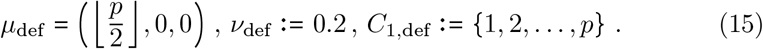

Having initialized the above parameters we follow the following pseudo code:

(I.) **for** *t* = 1, 2, …, *T* **do:**
  (1.) Draw a missingness scenario Ξ (^*t)*^ from the missingness parameters in (16), cf. Section 6.2.
  (2.) Draw mask **M**^(*t*)^ from Ξ^(*t*)^, cf. Section 6.2, and mask **X**_obs_ by **M**^(*t*)^ to obtain a new observed matrix 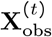.
  (3.) Set 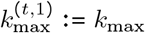and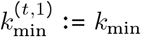.
  (4.) **for** *ℓ* = 1, 2, …, *b*_dp_ **do:**
    a. Compute

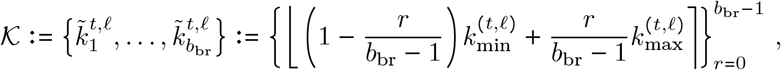

where ⌊·⌉ denotes the rounding to the next integer.
    b. **for** *s* = 1, 2, …, *b*_br_ **do:**
      i. Compute an imputed data matrix **X**_imp_ ^(*t*,*ℓ*,*s*)^ via k-nearest neighbour imputation with *k* set to 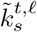 and with a time-out at *T*_time_ seconds.
      ii. if **X**_imp_^(*t*,*ℓ*,*s*)^ is computed successfully **do:** Round binary and count columns appropriately and compute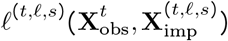 by (6.12) of the main text.
    c. Compute

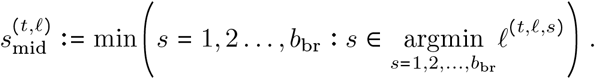
    d. Set

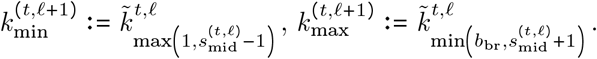
  (5.) Set

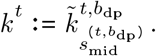
(II.) Compute *k* as the median of all { *k*^1^, *k*^(2)^, …, *k*^(*T*)^ }.

In the case that in the above pseudo code imputation is not successful (e.g. it runs out of time), the respective parameters are disregarded in the computation of *k*. In the case that all attempts at imputation fail for all 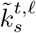 the parameter *k* is set to *k*_def_.

### C.2 MICE

To include MICE as a default MVI method in `ImputeBench` we use the R-package `mice`, see [91]. All parameters are set to default choices of `mice::mice`, except for `mincor` which is set to 0.1.

### C.3 missForest

To include missForest as a default MVI method in `ImputeBench` we use the R-package `missForest`, see [83]. All parameters are set to their respective defaults given in `missForest::missForest`.

### C.4 soft impute

To include soft impute as a default MVI method in `ImputeBench` we use the R-package `softImpute`, see [56]. All parameters but three are set to their respective defaults given in `softImpute::softImpute`. Regarding the three non-default parameter, in the following we detail the training of *λ* and how `rank.max` and `type` are set.

Given a data matrix **X**_obs_ ∈ (ℝ {*NA*}) ^*n*×*p*^ with missing entries. We fix

- `rank.max` is set to min *n, p* − 1.
- `type` is set to “svd”.

To train `lambda` of soft impute we start by defining the following parameters

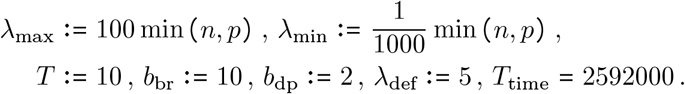

As when training the k-nearest neighbour imputation we define default missingness parameters

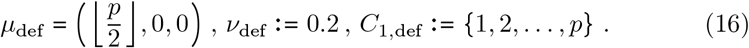

Having initialized the above parameters we follow the following pseudo code:

(I.) **for** *t* = 1, 2, …, *T* **do:**
  (1.) Draw a missingness scenario Ξ ^(*t)*^ from the missingness parameters in (16), cf. Section 6.2.
  (2.) Draw mask **M**^(*t*)^ from Ξ ^(*t*)^, cf. Section 6.2, and mask **X**_obs_ by **M**^(*t*)^ to obtain a new observed matrix 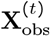.
  (3.) Set 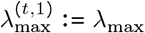and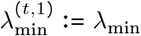.
  (4.) **for** *ℓ* = 1, 2, …, *b*_dp_ **do:**
    a. Compute

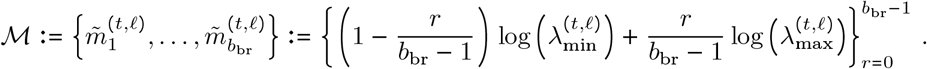
    b. Compute

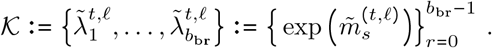
    c. **for** *s* = 1, 2, …, *b*_br_ **do:**
      i. Compute an imputed data matrix **X**_imp_ ^*t*,*ℓ*,*s*^ via soft impute with *λ* set to 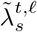 and with a time-out at *T*_time_ seconds.
      ii. if **X**_imp_^(*t*,*ℓ*,*s*)^ is computed successfully **do:** Round binary and count columns appropriately and compute 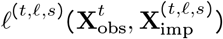 by (6.12) of the main text..
    d. Compute

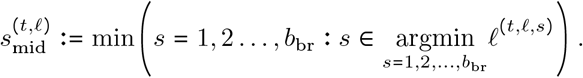
    e. Set

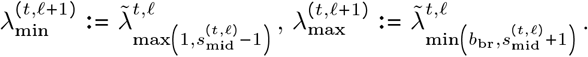
  (5.) Set

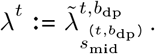
(II.) Compute *λ* as the median of all {*λ*^1^, *λ*^(2)^, …, *λ*^(*T*)^ }.

In the case that in the above pseudo code imputation is not successful (e.g. it runs out of time), the respective parameters are disregarded in the computation of *λ*. In the case that all attempts at imputation fail for all 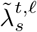 the parameter *λ* is set to *λ*_def_.

